# Emergence and spread of a SARS-CoV-2 variant through Europe in the summer of 2020

**DOI:** 10.1101/2020.10.25.20219063

**Authors:** Emma B. Hodcroft, Moira Zuber, Sarah Nadeau, Timothy G. Vaughan, Katharine H. D. Crawford, Christian L. Althaus, Martina L. Reichmuth, John E. Bowen, Alexandra C. Walls, Davide Corti, Jesse D. Bloom, David Veesler, David Mateo, Alberto Hernando, Iñaki Comas, Fernando González Candelas, SeqCOVID-SPAIN consortium, Tanja Stadler, Richard A. Neher

## Abstract

Following its emergence in late 2019, severe acute respiratory syndrome coronavirus 2 (SARS-CoV-2) has caused a global pandemic resulting in unprecedented efforts to reduce transmission and develop therapies and vaccines (WHO Emergency Committee, 2020; Zhu et al., 2020). Rapidly generated viral genome sequences have allowed the spread of the virus to be tracked via phylogenetic analysis (Worobey et al., 2020; Hadfield et al., 2018; Pybus et al., 2020). While the virus spread globally in early 2020 before borders closed, intercontinental travel has since been greatly reduced, allowing continent-specific variants to emerge. However, within Europe travel resumed in the summer of 2020, and the impact of this travel on the epidemic is not well understood. Here we report on a novel SARS-CoV-2 variant, 20E (EU1), that emerged in Spain in early summer, and subsequently spread to multiple locations in Europe. We find no evidence of increased transmissibility of this variant, but instead demonstrate how rising incidence in Spain, resumption of travel across Europe, and lack of effective screening and containment may explain the variant’s success. Despite travel restrictions and quarantine requirements, we estimate 20E (EU1) was introduced hundreds of times to countries across Europe by summertime travellers, likely undermining local efforts to keep SARS-CoV-2 cases low. Our results demonstrate how a variant can rapidly become dominant even in absence of a substantial transmission advantage in favorable epidemiological settings. Genomic surveillance is critical to understanding how travel can impact SARS-CoV-2 transmission, and thus for informing future containment strategies as travel resumes.

Severe acute respiratory syndrome coronavirus 2 (SARS-CoV-2) is the first pandemic where the spread of a viral pathogen has been globally tracked in near real-time using phylogenetic analysis of viral genome sequences (Worobey et al., 2020; Hadfield et al., 2018; Pybus et al., 2020). SARS-CoV-2 genomes continue to be generated at a rate far greater than for any other pathogen and more than 500,000 full genomes are available on GISAID as of February 2020 (Shu and McCauley, 2017).

In addition to tracking the viral spread, these genome sequences have been used to monitor mutations which might change the transmission, pathogenesis, or antigenic properties of the virus. One mutation in particular, D614G in the spike protein, has received much attention. This variant (Nextstrain clade 20A) seeded large outbreaks in Europe in early 2020 and subsequently dominated the outbreaks in the Americas, thereby largely replacing previously circulating lineages. This rapid rise led to the suggestion that this variant is more transmissible, which has since been corroborated by phylogenetic (Korber et al., 2020; Volz et al., 2020) and experimental evidence (Plante et al., 2020; Yurkovetskiy et al., 2020).

Following the global dissemination of SARS-CoV-2 in early 2020 (Worobey et al., 2020), intercontinental travel dropped dramatically. Within Europe, however, travel and in particular holiday travel resumed in summer (though at lower levels than in previous years) with largely uncharacterized effects on the pandemic. Here we report on a novel SARS-CoV-2 variant 20E (EU1) (`S:A222V`) that emerged in early summer 2020, presumably in Spain, and subsequently spread to multiple locations in Europe. Over the summer, it rose in frequency in parallel in multiple countries. As we report here, this variant, 20E (EU1), and a second variant 20A.EU2 with mutation `S477N` in the spike protein accounted for the majority of sequences in Europe in the autumn of 2020.

## Multiple variants emerged in Summer 2020 in Europe

Figure 1 shows a time scaled phylogeny of sequences sampled in Europe through the end of November and their global context, highlighting the variants in this manuscript. Clade 20A and its daughter clades 20B and 20C have variant `S:D614G` and are colored in yellow. A cluster of sequences in clade 20A has an additional mutation `S:A222V` colored in orange. We designate this cluster as 20E (EU1) (this cluster consists of lineage B.1.177 and its sublineages (Rambaut et al., 2020)).

**FIG. 1.**
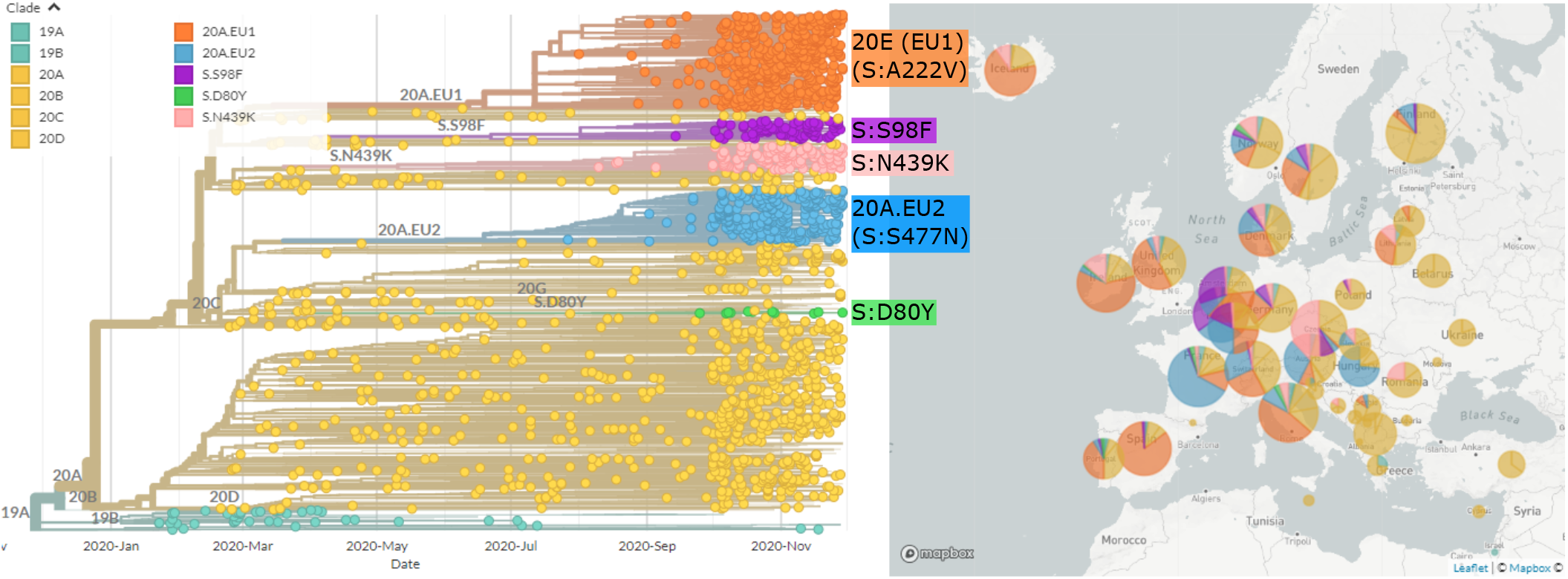
Phylogenetic overview of SARS-CoV-2 in Europe through the end of November. The tree shows a representative sample of isolates from Europe colored by clade and by the variants highlighted in this paper. A novel variant (orange; 20E (EU1)) with mutation `S:A222V` on a `S:D614G` background emerged in early summer and is common in most countries with recent sequences. A separate variant (20A.EU2, blue) with mutation `S:S477N` is prevalent in France. On the right, the proportion of sequences belonging to each variant (through the duration of the pandemic) is shown per country. Tree and visualization were generated using the Nextstrain platform (Hadfield et al., 2018) as described in methods.

In addition to the 20E (EU1) cluster we describe here, an additional variant (20A.EU2; blue in Fig. 1) with several amino acid substitutions, including `S:S477N` and mutations in the nucleocapsid protein, has become common in some European countries, particularly France (Fig. S6). The `S:S477N` substitution has arisen multiple times independently, for example in a variant in clade 20B that has dominated the outbreak in Oceania during the southern-hemisphere winter (now identified as 20F). The position 477 is close to the receptor binding site (Fig. S1). Residue `S477` is part of the epitope recognized by the S2E12 and C102 neutralizing antibodies (Barnes et al., 2020; Tortorici et al., 2020) and the detection of multiple variants at this position, such as `S477N`, might have resulted from the selective pressure exerted by the host immune response.

Several other smaller clusters defined by the spike mutations `D80Y`, `S98F`, `N439K` are also seen in multiple countries (see Table I and Fig. S6). While none of these have reached the prevalence of 20E (EU1) or 20A.EU2, some have attracted attention in their own right: `S:N439K` has appeared twice in the pandemic (Thomson et al., 2020) as well as numerous times independently. It is found across Europe (particularly Ireland, the UK, and Czech Republic), is located in the receptor binding domain (RBD), and reduces neutralization by antibody C135 (Barnes et al., 2020; Weisblum et al., 2020). Focal phylogenies for these, and other variants mentioned in this paper, as well as updated phylogenies of SARS-CoV-2 in Europe and individual European countries can be found at nextstrain.org/groups/neherlab. Further detailed analyses of the individual clusters discussed here are available at CoVariants.org.

**TABLE I.**
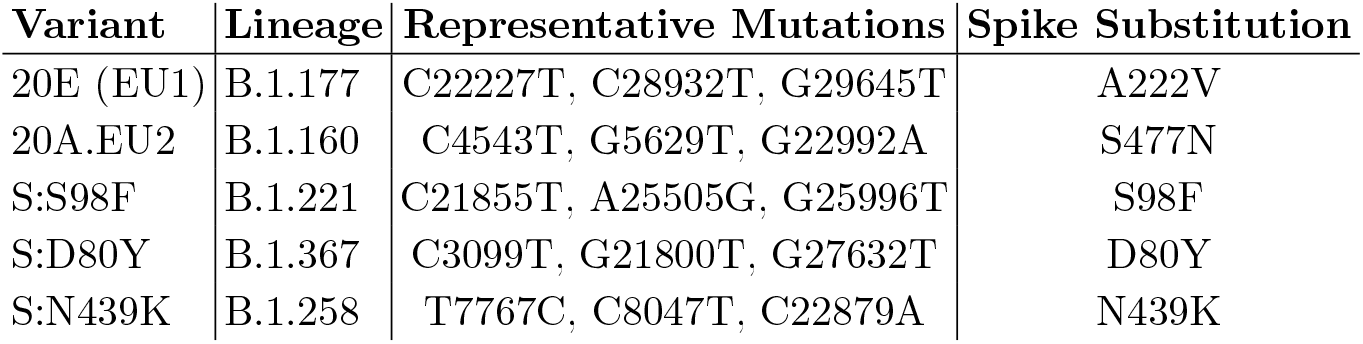
Representative mutations of 20E (EU1) (the focus of this study) and other notable variants. When a lineage definition matches the variant definition, it is given in column 2 (Rambaut et al., 2020).

### Antigenic and functional characterization of S:A222V

Our analysis here focuses on the variant 20E (EU1) with substitution `S:A222V`. `S:A222V` is in the spike protein’s domain A (Figure S1) also referred to as the N-terminal domain (NTD) (Tortorici et al., 2020; McCallum et al., 2020; Walls et al., 2020), which is not known to play a direct role in receptor binding or membrane fusion for SARS-CoV-2. However, mutations can sometimes mediate long-range effects on protein conformation or stability.

To evaluate if the A222V mutation affects the conformation of the SARS-CoV-2 spike glycoprotein, we probed binding of the benchmark COVID-19 convalescent patient plasma from the National Institute for Biologicals Standards and Control, two neutralizing monoclonal antibodies recognizing the RBD, namely S2E12 and S309 (Tortorici et al., 2020; Pinto et al., 2020; Walls et al., 2020) and a NTD-specific neutralizing monoclonal antibody (4A8) (Chi et al., 2020). The dose-response curves were indistinguishable for the SARS-CoV-2 2P S and the SARS-CoV-2 2P A222V D614G S ectodomain trimers, as observed by ELISA, Fig.S3a-d S3. These results are in agreement with a recent study showing that binding of several NTD-specific neutralizing antibodies were unaffected by the A222V mutation (McCallum et al., 2021). Collectively, these data indicate that the A222V substitution does not affect the SARS-CoV-2 S antigenicity appreciably.

To test whether the `S:A222V` mutation had an obvious functional effect on spike’s ability to mediate viral entry, we produced lentiviral particles pseudotyped with spike either containing or lacking the `A222V` mutation in the background of the `D614G` mutation and deletion of the end of spike’s cytoplasmic tail. Lentiviral particles with the `A222V` mutant spike had slightly higher titers than those without (mean 1.3-fold higher), although the difference was not statistically significant after normalization by p24 concentration (Fig. S2). Therefore, `A222V` does not lead to the same large increases in the titers of spike-pseudotyped lentivirus that has been observed for the `D614G` mutation (Korber et al., 2020; Yurkovetskiy et al., 2020), which is a mutation that is now generally considered to have increased the fitness of SARS-CoV-2 (Volz et al., 2020; Plante et al., 2020). However, we note that this small effect must be interpreted in equivocal terms, as the effects of mutations on actual viral transmission in humans are not always paralleled by measurements made in highly simplified experimental systems such as the one used here.

In addition to `S:A222V`, 20E (EU1) has the amino acid mutations `ORF10:V30L`, `N:A220V` and `ORF14:L67F`. However, there is little evidence of the functional relevance of `ORF10` and `ORF14` (Pancer et al., 2020; Finkel et al., 2020). Different mutations at positions 180 and 220 in `N` are observed in almost every major lineage and we are not aware of any evidence suggesting that these mutations have important phenotypic consequence. Therefore, we examined epidemiological and evolutionary evidence to assess if the variant showed evidence of enhanced transmissibilty in humans.

### Early observations of 20E (EU1)

The earliest sequences identified date from the 20th of June, when 7 Spanish sequences and 1 Dutch sequence were sampled. The next non-Spanish sequence was from the UK (Wales) on the 7th July, with a Belgian sequence sampled on the 17th and a Swiss sampled on the 22nd. By the end of July, samples from Spain, the Netherlands, the UK (England, Northern Ireland, Wales), Switzerland, Ireland, Belgium, and Norway were identified as being part of the cluster. By the 22nd August, the cluster also included sequences from France, Denmark, more of the UK (Scotland), Germany, Latvia, Sweden, and Italy. Four sequences from Hong Kong, 17 from Australia, 27 from New Zealand, and 8 sequences from Singapore, presumably exports from Europe, were first detected in mid-August (Hong Kong, Australia), mid-September (New Zealand), and mid-October (Singapore).

The proportion of sequences from several countries which fall into 20E (EU1), by ISO week, is plotted in Fig. 2. 20E (EU1) first rose in frequency in Spain, jumping to around 50% prevalence within a month of the first sequence being detected before rising to 80%. In many countries, including the United Kingdom, France, Ireland, Denmark, and Switzerland we observe a gradual rise starting in mid-July before settling at a level between 15 and 80% in September or October. In contrast, Norway observed a sharp peak in early August, but seemed to bring 20E (EU1)cases down quickly, though they began growing again in September. The date ranges and number of sequences observed in this cluster are summarized in Table SI.

**FIG. 2.**
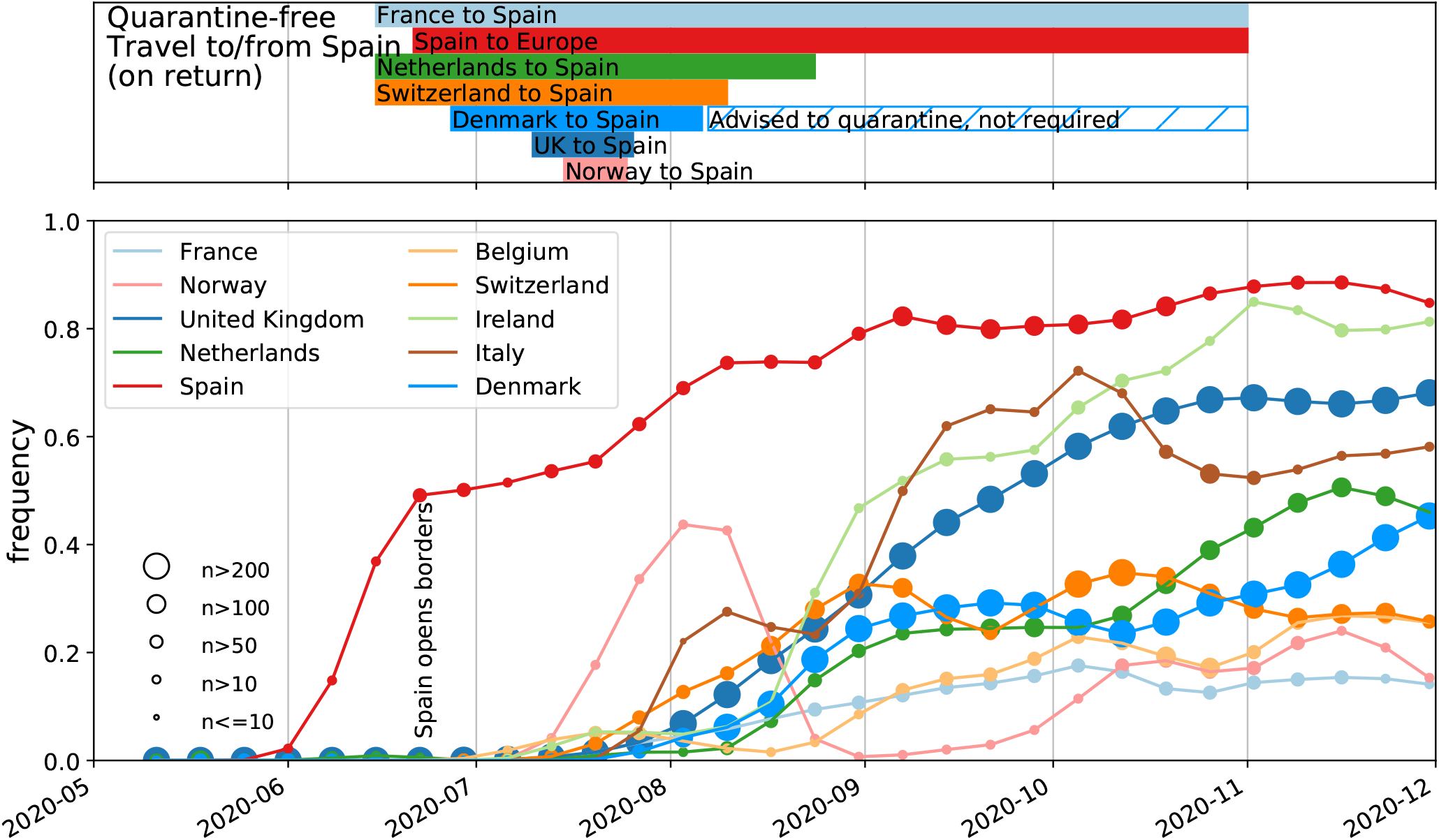
Frequency of submitted samples are 20E (EU1) in selected countries, with quarantine-free travel dates shown above. We include the eight countries which have at least 200 sequences from 20E (EU1), as well as Norway and France, to illustrate points in the text. The symbol size indicates the number of available sequence by country and time point in a non-linear manner. Travel restrictions from selected countries are shown to/from Spain, as this is the probable origin of the cluster. Most European countries allowed quarantine-free travel to other (non-Spanish) countries in Europe for a longer period. When the last data point included only very few sequences, it has been dropped for clarity. Frequencies are smoothing using a Gaussian with *σ* = 1*w*.

### Initial expansion and spread across Europe

Fig. 3 shows a phylogeny based on data from samples collected before 2020-09-30 and available on GISAID in Jan 2021, as described in Methods. The phylogeny is collapsed to group diversity possibly stemming from within-country transmission into sectors of the pie-charts (see Fig. S12) for selected countries. The tree indicates that 20E (EU1) harbors substantial diversity and most major genotypes within the 20E (EU1) cluster have been observed in many European countries. Since it is unlikely that phylogenetic patterns that are sampled in multiple countries arose independently, it is reasonable to assume that the majority of mutations observed in the tree arose once and were carried (possibly multiple times) between countries. We use this rationale to provide lower bounds on the number of introductions to different countries. Throughout July and August 2020, Spain had a higher per capita incidence than most other European countries (see Fig S4) and 20E (EU1) was much more prevalent in Spain then elsewhere, suggesting Spain as likely origin of most 20E (EU1) introductions to other countries. The first sequences in 20E (EU1) were sampled on the 20th of June in Spain and the Netherlands. This Dutch sequence nests within the diversity of early sequences from Spain, suggesting this sequence is the result of the earliest sampled export of the variant outside of Spain, consistent with and early uptick of travel from Spain to the Netherlands (Fig. 4 A). Other sequences in 20E (EU1) that are dated prior to June 2020 have implausible phylogenetic positions and are likely mis-dated (these are excluded, see Methods).

**FIG. 3.**
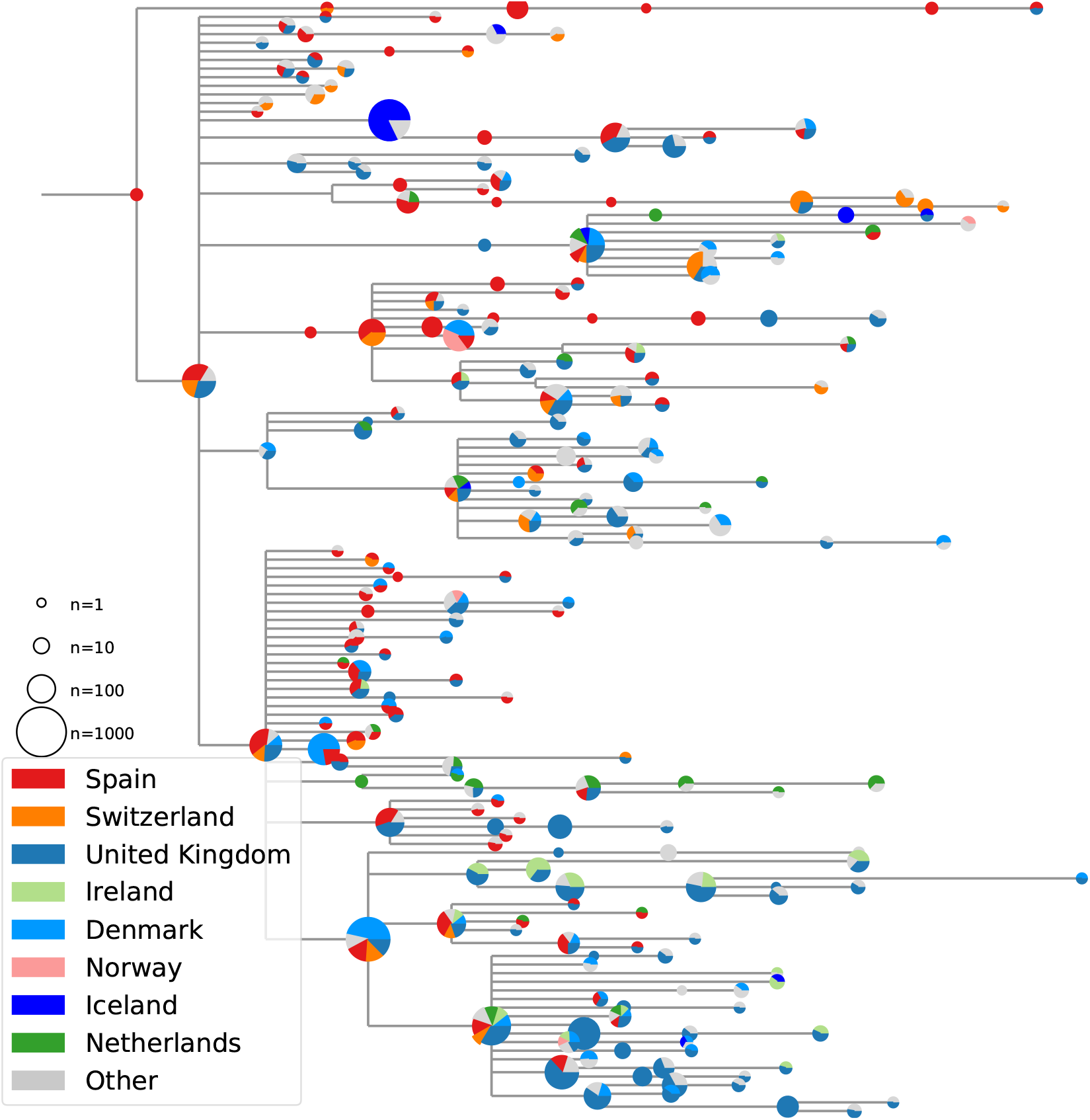
Collapsed genotype phylogeny. The phylogeny shown is the subtree of the 20E (EU1) cluster using data from samples collected before 30 Sept 2020 and available on GISAID as of Jan 2021, with sequences carrying all six defining mutations. Pie charts show the representation of sequences from selected countries at each node. Size of the pie chart indicates the total number of sequences at each node. Pie chart fractions scale non-linearly with the true counts (fourth root) to ensure all countries are visible and branch lengths are jittered to reduce overlap.

**FIG. 4.**
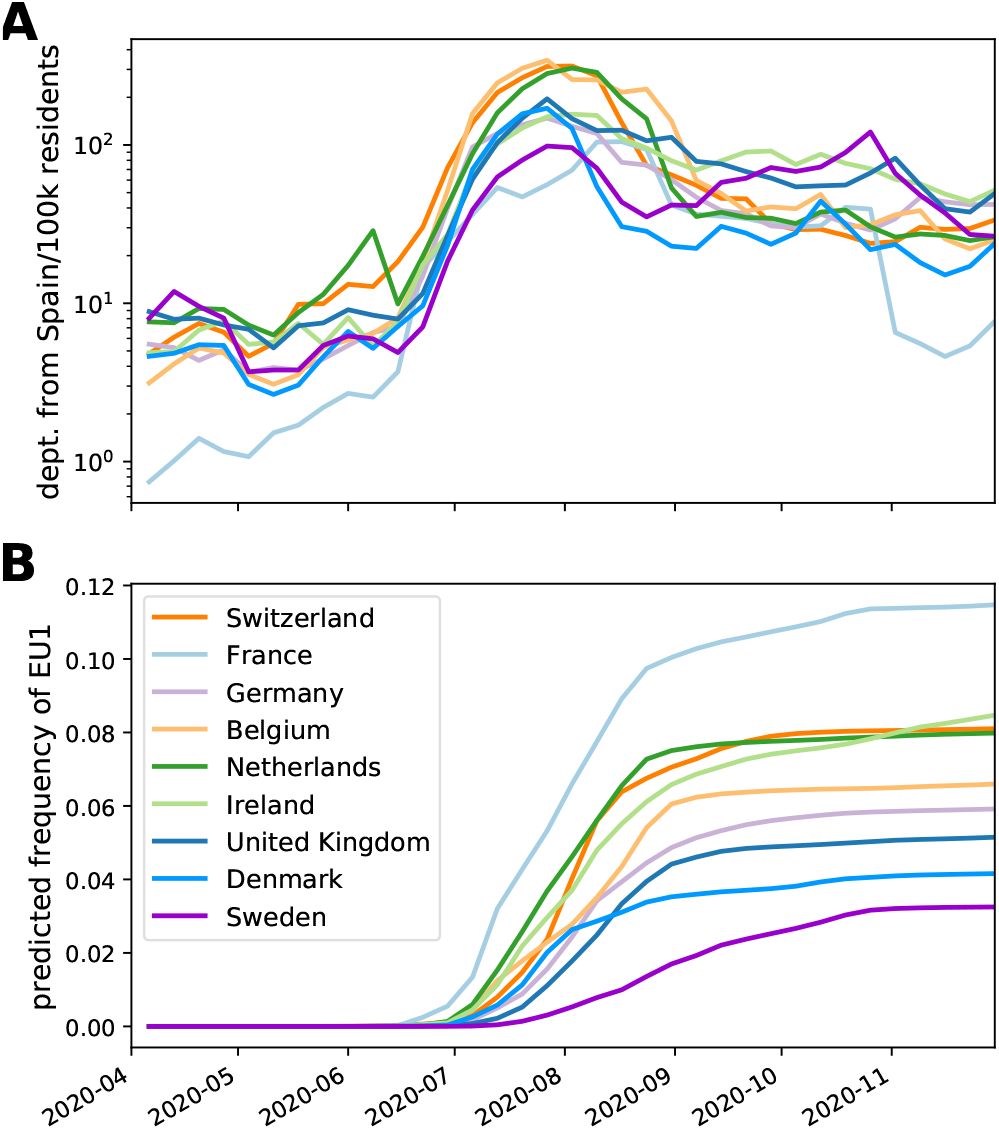
Travel volume and contribution of imported infections. Travel from Spain to other European countries resumed in July (though low compared to previous years). Assuming that travel returnees are infected at the average incidence of Spanish province they visited and transmit the virus at the rate of their resident population, imports from Spain are expected to account between 2 and 12% of SARS-CoV-2 cases after the summer. Traveler incidence is calculated using case and travel data at the level of provinces. Note that this model only accounts for contribution of summer travel and that stochastic fluctuations and other variants after the summer will results in further variation in the frequency of 20E (EU1). See Methods and Fig. S8.

Epidemiological data from Spain indicates the earliest sequences in the cluster are associated with two known outbreaks in the north-east of the country. The variant seems to have initially spread among agricultural workers in Aragon and Catalonia, then moved into the local population, where it was able to travel to the Valencia Region and on to the rest of the country (though sequence availability varies between regions). This initial expansion may have been critical in increasing the cluster’s prevalence in Spain just before borders re-opened.

Most basal genotypes have been observed both in Spain and a remarkably large number of other countries, suggesting repeated exports. However, the 801 sequences from Spain contributing to Fig. 3 likely do not represent the full diversity. Variants found only outside of Spain may reflect diversity that arose in secondary countries, or may represent diversity not sampled in Spain. In particular, as the UK sequences much more than any other country in Europe, it is not unlikely they may have sampled diversity that exists in Spain or elsewhere but has only been sampled in the UK. Despite limitations in sampling, Fig. 3 clearly shows that most major genotypes in this cluster were distributed to multiple countries, suggesting that many countries have experienced multiple introductions of identical genotypes that cannot be fully identified from the phylogeny. This is consistent with the large number of introductions estimated from travel data, discussed below. While initial introductions of the variant likely originated from Spain, 20E (EU1) cases outside of Spain surpassed those in Spain in late September and later cross-border transmissions likely originated in other European countries (see Fig S4B and 20E (EU1) Nextstrain build online). In the supplementary text we provide a brief discussion of travel restrictions and measures imposed by selected European countries and the associated patterns of 20E (EU1) introductions.

Fig. S5 shows the distribution of sequence clusters compatible with onward transmission within countries outside of Spain, highlighting two different patterns. Norway and Iceland, for example, seem to have experienced only a small number of introductions over the summer that led to substantial further spread. In Fig. 3, the majority of sequences from these countries fall into one sector, the remainder are singletons or very small clusters that have not spread. Unlike the initial introduction, later sequences in Norway or Iceland often cluster more closely with diversity in European countries other than Spain, which may suggest further introductions came from third countries (see 20E (EU1) Nextstrain build online).

In contrast, countries like Switzerland, the Netherlands, or the United Kingdom have sampled sequences that correspond to a large number of independent introductions that include most major and many minor genotypes observed in Spain. Many sequences sampled in Ireland are closely associated with sequences sampled in the UK, which might indicated exchange with the UK or shared holiday destinations. As described in the supplementary text, Ireland never allowed quarantine-free travel to Spain, but travel figures (Fig 4 A) suggest it was a popular destination nonetheless.

### No evidence for transmission advantage of 20E (EU1)

During a dynamic outbreak, it is particularly difficult to unambiguously tell whether a particular variant is increasing in frequency because it has an intrinsic advantage, or because of epidemiological factors (Grubaugh et al., 2020). In fact, it is a tautology that every novel big cluster must have grown recently and multiple lines of independent evidence are required in support of an intrinsically elevated transmission potential.

The cluster we describe here – 20E (EU1) (`S:A222V`) – was dispersed across Europe initially mainly by travelers to and from Spain. Many EU and Schengen-area countries, including Switzerland, the Netherlands, and France, opened their borders to other countries in the bloc on 15th June. Travel resumed quickly and peaked during July and August, see Fig. 4. The number of confirmed SARS-CoV-2 cases in Spain rose from around 10 cases per 100k inhabitants per week in early July to 100 in late August, while case number remained low in most of Europe during this time. To explore whether repeated imports are sufficient to explain the rapid rise in frequency and the displacement of other variants, we first estimated the number of expected introductions of 20E (EU1) based on the number of visitors from a particular country to different provinces of Spain and the SARS-CoV-2 incidence in the provinces. Taking reported incidence in the provinces at face value and assuming that returning tourists have a similar incidence, we expect 380 introductions of 20E (EU1) into the UK over the summer (6 July-27 Sept, see Table SII and Fig. 4 for tourism summaries (Instituto Nacional de Estadistica, 2020) and departure statistics (Aena.es, 2020). Similarly, for Germany and Switzerland we would expect around 320 and 90 introductions of 20E (EU1), respectively. We then create a simple model that also incorporates the incidence in the country where travellers are returning to and onward spread of imported 20E (EU1) cases to estimate the frequency of 20E (EU1) in countries across Europe over time (see Fig. 4). This model assumes that 20E (EU1) spread at the same rate as other variants in the resident countries and predicts that the frequencies of 20E (EU1) would start rising in July, continue to rise through August, and be stable thereafter in concordance with observations in many countries including Switzerland, Denmark, France, Germany or the Netherlands (see Fig. 4 B).

While the shape of the expected frequency trajectories from imports in Fig. 4 B is consistent with observations, this naive import model underestimates the final observed frequency of 20E (EU1) by between 1- and 11-fold depending on the country, see Fig. S9. This discrepancy might be to due to either intrinsically faster transmission of 20E (EU1) or due to underestimation of introductions. Underestimates might be due to country-specific reporting and behavioural factors such as the relative ascertainment rate in source and destination populations and the fact that risk of exposure and onward transmission are likely increased by travel-related activities both abroad, en route, and at home. Furthermore, SARS-CoV-2 incidence in holiday destinations may not be well-represented by the provincial averages used in the model. For example, during the first wave in spring, some ski resorts had exceptionally high incidence and contributed disproportionately to dispersal of SARS-CoV-2 (Knabl et al., 2020; Correa-Martinez et al., 2020). The fact that the rapid increase of the frequency of 20E (EU1) slowed or stopped in most countries after the summer travel period and didn’t fully replace other variants is consistent with import driven dynamics with little or no competitive advantage.

The notion that an underestimated incidence in travel returnees rather than faster spread of 20E (EU1) is the major contributor to above discrepancy is supported by the fact that German authorities report about 2.2 times as many cases with suspected infection in Spain than the model predicts (982 reported vs 452 estimated from 6 July-13 Sept regardless of variant), see Fig. S10 A. Switzerland reported 131 infections in travel returnees, while the model predicts 130. After adjusting imports for the 37% of Swiss case reports without exposure information, the model underestimates introductions 1.6-fold. Taken together, these observations suggest that underestimation of incidence in travelers can explain much of the discrepancy between the predicted and observed frequency dynamics of 20E (EU1). Countries with small (1-4 fold) and large (8-11 fold) discrepancies tend to visit distinct destinations in Spain, see Figs. S9 and S8, suggesting variation in behavior and incidence in the holiday destination may explain some of the underestimation.

To investigate the possibility of faster growth of 20E (EU1) introductions, we identified 20E (EU1) and non-20E (EU1) introductions into Switzerland and their downstream Swiss transmission chains in the available sequence data. These data suggest 34 or 291 introductions of the 20E (EU1) variant depending on the criterion used to assign sequences to putative transmission chains (see Methods). Phylodynamic estimates of the effective reproductive number (*R*_*e*_) through time for introductions of 20E (EU1) and for other variants (see Fig. S11) suggest a tendency for 20E (EU1) introductions to transiently grow faster. This transient signal of faster growth, however, is more readily explained with behavioral differences and increased travel-associated transmission than intrinsic differences to the virus. We repeated the phylodynamic analysis with a pan-European set of putative introductions showing similar patterns as observed for Switzerland.

These patterns are further consistent with the fact that Swiss cases with likely exposure in Spain tended to be in younger individuals (median 30 years, IQR 23-42.25 years) than cases acquired in Switzerland (median 35 years, IQR 24-51 years). These younger individuals tend to have more contacts than older age groups (Mossong et al., 2008; Jarvis et al., 2020). Such association with particular demographics will decay rapidly and with it any associated increased transmission as inferred by phylodynamics.

Most 20E (EU1) introductions are expected to have occurred towards the end of summer when incidence in Spain was rising and return travel volume peaked. Comparatively high incidence of non-20E (EU1) variants at this time and hence a relatively low impact of imported variants (e.g. Belgium, see Fig. S4) might explain why 20E (EU1) remains at low frequencies in some countries despite high-volume travel to Spain. To date, 20E (EU1) has not been observed in Russia, consistent with little travel to/from Spain and continuously high SARS-CoV-2 incidence (Instituto Nacional de Estadistica, 2020).

While we can not rule out that 20E (EU1) has a slight transmission advantage compared to other variants circulating at the time, most of its spread is explained by epidemiological factors, despite the fact that case numbers across Europe started to rise rapidly around the same time the 20E (EU1) variant started to become prevalent in multiple countries, (Fig. S4). However, countries where 20E (EU1) is rare (Belgium, France, Czech Republic - see Fig. S6) have seen similarly rapid increases, suggesting that this rise was not driven by any particular lineage and that 20E (EU1) has no difference in transmissibility. Furthermore, we observe in Switzerland that *R*_*e*_ increased in fall by a comparable amount for the 20E (EU1) and non-20E (EU1) variants (see (Fig. S11). The arrival of fall and seasonal factors are a more plausible explanation for the resurgence of cases (Neher et al., 2020).

## DISCUSSION

The rapid spread of 20E (EU1) and other variants underscores the importance of a coordinated and systematic sequencing effort to detect, track, and analyze emerging SARS-CoV-2 variants. This becomes even more urgent with the recent detection of several Variants of Concern (VoC) (Rambaut et al., 2020; Tegally et al., 2020; Faria et al., 2021). It is only through multi-country genomic surveillance that it has been possible to detect and track 20E (EU1) and other variants. However, genomic surveillance still varies greatly between countries, with variants more likely to be detected in countries that sequence a significant fraction of all cases, like the United Kingdom and Denmark in Europe.

When a new variant is observed, policy makers need a rapid assessment of whether the new variant increases the transmissibility of the virus, evades pre-existing immunity or has different clinical properties (Lauring and Hodcroft, 2021). In case of 20E (EU1) none of these properties seem to have changed substantially, making it an important example of how travel combined with large regional differences in prevalence can lead to substantial rapid shifts in the variant distribution without a dramatic transmission advantage. Such shifts that are driven predominantly by epidemiological factors are more likely in a low incidence setting, where a large fraction of cases can be due to introductions. In contrast the VoC 501Y.V1/B.1.1.7 spread across Europe in late 2020 while most countries, including the UK, where it first rose to prominence, reported high incidence. In such a high incidence setting, travel alone can not explain a rapid rise in frequency and the dynamics points to a bona fide transmission advantage. In depth characterization of a spectrum of such dynamics (no substantial advantage in case of 20E (EU1), moderate advantage in case of D614G (Volz et al., 2020), and a strong transmission advantage of 501Y.V1/B.1.1.7 (Davies et al., 2020) and 501Y.V2 (Pearson et al., 2021)) will facilitate assessment of emerging variants in the future.

Along similar lines, it is imperative to understand whether novel variants impact the severity of the disease. So far, we have no evidence for any such effect: the low mortality over the summer in Europe was predominantly explained by a much better ascertainment rate and a marked shift in the age distribution of confirmed cases. This variant was not yet prevalent enough in July and August to have had a big effect. As sequences and clinical outcomes for patients infected with this variant become available, it will be possible to better infer whether this lineage has any impact on disease prognosis. Finally, our analysis highlights that countries should carefully consider their approach to travel when largescale inter-country movement resumes across Europe. We show that holiday travel in summer 2020 resulted in unexpectedly high levels of introductions and onward spread across Europe. Whether the 20E (EU1) variant described here has rapidly spread due to a transmission advantage or due to epidemiological factors alone, its repeated introduction and rise in prevalence in multiple countries implies that the summer travel guidelines and restrictions were generally not sufficient to prevent onward transmission of introductions. Travel precautions such as quarantine should in principle have prevented spread of SARS-CoV-2 infections acquired abroad, but in practice failed to have the desired effect. While long-term travel restrictions and border closures are not tenable or desirable, identifying better ways to reduce the risk of introducing variants, and ensuring that those which are introduced do not go on to spread widely, will help countries maintain often hard-won low levels of SARS-CoV-2 transmission.

## Supporting information

Acknowledgement Table

## Data Availability

All sequence data is available on GISAID (gisaid.org), and the code for the analyses performed is available on Github and linked in the manuscript text.

https://nextstrain.org/groups/neherlab/ncov/20A.EU1

https://github.com/hodcroftlab/covariants/tree/eu1_paper

## Acknowledgements

We are grateful to researchers, clinicians, and public health authorities for making SARS-CoV-2 sequence data available in a timely manner. We also wish to thank the COVID-19 Genomics UK consortium for their notable sequencing efforts, which have provided more than half of the sequences currently publicly available. We would also like to thank the Swiss Federal Office of Public Health (FOPH) for providing access to their data. This work was supported by the Swiss National Science Foundation (SNSF) through grant numbers 31CA30 196046 (to RAN, EBH, CLA), 31CA30 196267 (to TS), European Union’s Horizon 2020 research and innovation programme - project EpiPose (No 101003688) (MLR, CLA), core funding by the University of Basel and ETH Zürich, the National Institute of General Medical Sciences (R01GM120553 to DV), the National Institute of Allergy and Infectious Diseases (DP1AI158186 and HHSN272201700059C to DV), a Pew Biomedical Scholars Award (DV), an Investigators in the Pathogenesis of Infectious Disease Awards from the Burroughs Wellcome Fund (DV and JDB), a Fast Grants (DV), and NIAID grants R01AI141707 (JDB) and F30AI149928 (KHDC). SeqCOVID-SPAIN is funded by the Instituto de Salud Carlos III project COV20/00140, Spanish National Research Council and ERC StG 638553 to IC and BFU2017-89594R from MICIN to FGC. JDB is an Investigator of the Howard Hughes Medical Institute.

## Transparency declaration

DV is a consultant for Vir Biotechnology Inc. DC is an employee of Vir Biotechnology and may hold shares in Vir Biotechnology. The Veesler laboratory has received an unrelated sponsored research agreement from Vir Biotechnology Inc. AH is a co-founder of Kido Dynamics, DM is employed by Kido Dynamics. The other authors declare no competing interests.

## Authors’ contribution

EBH identified the cluster, led the analysis, created figures, and drafted the manuscript. RAN analysed data, created figures, and drafted the manuscript. MZ, SN, TGV, CLA, TS, and MLR analysed data and created figures. VD and ACW investigated structural aspects and created figures. JDB, JEB, ACW, DC, and KHDC performed experimental assays and created figures. IC and FGC interpreted the origin of the cluster and contributed data. DM and AH contributed and interpreted data. All authors contributed to and approved the final manuscript.

## METHODS

### Phylogenetic analysis

We use the Nextstrain pipeline for our phylogenetic analyses https://github.com/nextstrain/ncov/ (Hadfield et al., 2018). Briefly, we align sequences using mafft (Katoh et al., 2002), subsample sequences (see below), add sequences from the rest of the world for phylogenetic context based on genomic proximity, reconstruct a phylogeny using IQ-Tree (Minh et al., 2019) and infer a time scaled phylogeny using TreeTime (Sagulenko et al., 2018). For computational feasibility, ease of interpretation, and to balance disparate sampling efforts between countries, the Nextstrain-maintained runs sub-sample the available genomes across time and geography, resulting in final builds of ∼5,000 genomes each. After sub-sampling, the 20E (EU1) cluster within the Nextstrain build contains 5,145 sequences, 3,369 of which are unique (accounting for `N`s).

Sequences were downloaded from GISAID at the end of January and analysed using the nextstrain/ncov workflow, using a cutoff date of the 30 Sept (Fig 3) or 30 Nov (all other analyses). These dates were chosen to focus first on the introductions over the summer (for 30 Sept) and then to highlight ongoing circulation through the autumn (30 Nov) prior to the spread of the variants of concern identified in December 2020 and January 2021. A table acknowledging the invaluable contributions by many labs is available as a supplement. The Swiss SARS-CoV-2 sequencing efforts are described in (Nadeau et al., 2020) and (Stange et al., 2020). The majority of Swiss sequences used here are from the Nadeau et al. (2020) data set, the remainder are available on GISAID.

### Defining the 20E (EU1) Cluster

The cluster was initially identified as a monophyletic group of sequences stemming from the larger 20A clade with amino acid substitutions at positions `S:A222V`, `ORF10:V30L`, and `N:A220V` or `ORF14:L67F` (overlapping reading frame with N), corresponding to nucleotide mutations `C22227T`, `C28932T`, and `G29645T`. In addition, sequences in 20E (EU1) differ from their ancestors by the synonymous mutations `T445C`, `C6286T`, and `C26801G`.

The sub-sampling of the standard Nextstrain analysis means that we are not able to visualise the true size or phylogenetic structure of the cluster in question. To specifically analyze this cluster using almost all available sequences, we designed a specialized build which focuses on cluster-associated sequences and their most genetically similar neighbours. For computational reasons, we limit the number of samples to 900 per country per month. As only the UK has more sequences than this per month, this results in a random downsampling of sequences from the UK for the months of August, September, and October. Further, we excluded several problematic sequences due to high intra-sample variation, wrong dates, and over-divergence (divergence values are implausible given the provided dates). A full list of the sequences excluded (and the reason why) is given on github in bad sequences.py.

We identify sequences in the cluster based on the presence of nucleotide substitutions at positions 22227, 28932, and 29645 and use this set as a ‘focal’ sample in the nextstrain/ncov pipeline. This selection will exclude any sequences with no coverage or reversions at these positions, but the similarity-based sampling during the Nextstrain run will identify these, as well as any other nearby sequences, and incorporate them into the dataset. We used these three mutations as they included the largest number of sequences that are distinct to the cluster. By this criterion, there are currently 60,316 sequences in the cluster sampled before 30 November 2020. To visualise the changing prevalence of the cluster over time, we plotted the proportion of sequences identified by the four substitutions described above as a fraction of the total number of sequences submitted, per ISO week. Frequencies of other clusters are identified in an analogous way.

### Phylogeny and Geographic Distribution

The size of the cluster and number of unique mutations among individual sequences means that interpreting overall patterns and connections between countries is not straightforward. We aimed to create a simplified version of the tree that focuses on connections between countries and de-emphasizes onward transmissions within a country. As our focal build contains ‘background’ sequences that do not fall within the cluster, we used only the monophyletic clade containing the four amino-acid changes and three synonymous nucleotide changes that identify the cluster. Then, subtrees that only contain sequences from one country were collapsed into the parent node. The resulting phylogeny contains only mixed-country nodes and single-country nodes that have mixed-country nodes as children. (An illustrative example of this collapsing can be seen in Fig. S12.) Nodes in this tree thus represent ancestral genotypes of subtrees: sequences represented within a node may have further diversified within their country, but share a set of common mutations. We count all sequences in the subtrees towards the geographic distribution represented in the pie-charts in Fig. 3.

This tree allows us to infer lower bounds for the number of introductions to each country, and to identify plausible origins of those introductions. It is important to remember that, particularly for countries other than the United Kingdom, the full circulating diversity of the variant is probably not being captured, thus intermediate transmissions cannot be ruled out. In particular, the closest relative of a particular sequence will often have been sampled in the UK simply because sequencing efforts in the UK exceed most other countries by orders of magnitude. It is, however, not our goal to identify all introductions but to investigate large scale patterns of spread in Europe.

### Travel volume and destination

Mobile phone roaming data were used to estimate the number of visitors from a given country departing from a given province for each calendar week. The mobile phone record data set contains approximately 13 million devices, with over 2.6 million roamers. A visitor is considered to be departing the country during a given week if they are not seen in the data set for the next eight weeks. The nationality of a visitor is inferred from the Mobile Country Code (MCC). The total number of unique visitors is aggregated for each province and each week in the period of study; these totals are then scaled using official statistics as reference to account for the partial coverage of data set.

### Estimation of contributions from imports

To estimate how the frequency of 20E (EU1) is expected to change in country *X* due to travel, we consider the following simple model: A fraction *α*_*i*_ of the population of *X* returns from Spain every week *i* (estimated from roaming data, see above) and is infected with 20E (EU1) with a probability *p*_*i*_ given by its per capita weekly incidence in Spain. Incidence is the weighted average over incidence in Spanish provinces by the distribution of visitors across the provinces. The week-over-week fold change of the epidemic within *X* is calculated as *g*_*i*_ = (*c*_*i*_ − *α*_*i*_*p*_*i*_)*/c*_*i*−1_, where *c*_*i*_ is the per capita incidence in week *i* in *X*. This fold-change captures the local growth of the epidemic in country *X*. The total number of 20E (EU1) cases *v*_*i*_ in week *i* is hence *v*_*i*_ = *g*_*i*_*v*_*i*−1_ + *p*_*i*_*α*_*i*_, while the total number of non-20E (EU1) cases is *r*_*i*_ = *g*_*i*_*r*_*i*−1_. Running this recursion from mid-June to November results in the frequency trajectories in Fig. 4.

From 1 June 2020 to 30 September 2020, the Swiss Federal Office of Public Health (FOPH) reported 23,199 confirmed SARS-CoV-2 cases. 14,583 (62.9%) cases provided information about their likely place of exposure and country of infection in a clinical registration form. Of these, 3,304 (22.7%) reported an exposure abroad and 136 (0.9%) named Spain as the country of infection. The Robert-Koch-Institute reported statistics on likely country of infection by calendar week in their daily situation reports (Robert-Koch-Institute, 2020).

### Phylodynamic analysis of Swiss transmission chains

We identified introductions into Switzerland and downstream Swiss transmission chains by considering a tree of all available Swiss sequences combined with foreign sequences with high similarity to Swiss sequences (full procedure described in Nadeau et al. (2020)). Putative transmission chains were defined as majority Swiss clades allowing for at most 3 “exports” to third countries. Identification of transmission chains is complicated by polytomies in SARS-CoV-2 phylogenies and we bounded the resulting uncertainty by either (i) considering all sub-strees descending from the polytomy as separate introductions (called ‘max’ in Fig S11) and (ii) aggregating all into a single introduction (called ‘min’), see (Nadeau et al., 2020) for details. We further extended this analysis to include a pan-European dataset consisting of putative transmission chains defined via the collapsed phylogenies discussed earlier in the methods. Specifically, each section of a pie graph, which corresponds to a country-specific collection of sequences, was taken as a single introduction. Non-20E (EU1) *R*_*e*_ estimates were obtained from case data and the estimated frequency of 20E (EU1) in different countries.

The phylodynamic analysis of the transmission chains was performed using BEAST2 with a birth-death-model tree prior (Bouckaert et al., 2019; Stadler et al., 2013). 20E (EU1) and non-20E (EU1) variants share a sampling probability and log *R*_*e*_ has an Ornstein-Uhlenbeck prior, see Nadeau et al. (2020) for details (but note a different smoothing prior is used there).

### Enzyme-linked immunosorbent assay (ELISA)

384-well Maxisorp plates (Thermo Fisher) were coated overnight at room temperature with 3 *µ*g/mL in 20mM Tris pH 8 and 150mM NaCl of SARS-CoV-2 S2P (Pallenson et al 2017) or SARS-CoV-2 A222V-D614G S2P, produced as previously described in Walls et al. (2020). Briefly, Expi293F cells were transiently transcribed with a plasmid containing the spike protein and supernatant was clarified six days later prior to Ni Sepharose resin purification and flash freezing. Plates were slapped dry and blocked with Blocker Casein in TBS (Thermo Fisher) for one hour at 37°C. Plates were slapped dry and S2E12 (Tortorici et al., 2020) or S309 (Pinto et al., 2020) antibodies were serially diluted 1:3 with a starting concentration of 1000nM in TBST or NIBSC human plasma (20/130 https://www.nibsc.org/documents/ifu/20-130.pdf) was serially diluted 1:3 starting at 1:4 of original concentration in TBST and added to the plate for one hour at 37°C. Plates were washed 4x with TBST using a 405 TS Microplate Washer (BioTek) followed by addition of 1:5,000 goat anti-human Fc IgG-HRP (Thermo Fisher) for one hour at 37°C. Plates were washed 4x and TMB Microwell Peroxidase (Seracare) was added. The reaction was quenched after 1-2 minutes with 1 N HCl and the A450 of each well was read using a Varioskan Lux plate reader (Thermo Fisher).

### Pseudotyped Lentivirus Production and Titering

The `S:A222V` mutation was introduced into the protein-expression plasmid HDM-Spike-d21-D614G, which encodes a codon-optimized spike from Wuhan-Hu-1 (Genbank NC 045512) with a 21-amino acid cytoplasmic tail deletion and the D614G mutation (Greaney et al., 2020). This plasmid is also available on AddGene (plasmid 158762). We made two different versions of the `A222V mutant` that differed only in which codon was used to introduce the valine mutation (either `GTT` or `GTC`). The sequences of these plasmids (HDM Spike-d21-D614G-A222V-GTT and HDM Spike-d21-D614G-A222V-GTC) are available as supplement files at github.com/emmahodcroft/cluster scripts/.

Spike-pseudotyped lentiviruses were produced as described in (Crawford et al., 2020). Two separate plas-mid preps of the `A222V` (`GTT`) spike and one plasmid prep of the `A222V` (`GTC`) spike were each used in duplicate to produce six replicates of `A222V` spike-pseudotyped lentiviruses. Three plasmid preps of the initial `D614G` spike plasmid (with the 21-amino acid cytoplasmic tail truncation) were each used once used to make three replicates of `D614G`spike-pseudotyped lentiviruses. All viruses were titered in duplicate.

Lentiviruses were produced with both Luciferase IRES ZsGreen and ZsGreen only lentiviral backbones (Crawford et al., 2020), and then titered using luciferase signal or percentage of fluorescent cells, respectively. All viruses were titered in 293T-ACE2 cells (BEI NR-52511) as described in (Crawford et al., 2020), with the following modifications. Viruses containing luciferase were titered starting at a 1:10 dilution followed by 5 serial 2-fold dilutions. The Promega BrightGlo luciferase system was used to measure relative luciferase units (RLUs) ∼ 65 hours post-infection and RLUs per mL were calculated at each dilution then averaged across all dilutions for each virus. Viruses containing only ZsGreen were titered starting at a 1:3 dilution followed by 4 serial 5-fold dilutions. The 1:375 dilution was visually determined to be ∼1% positive about 65 hours post-infection and was used to calculate the percent of infected cells using flow cytometry (BD FACSCelesta cell analyzer). Viral titers were then calculated using the percentage of green cells via the Poisson formula. To normalize viral titers by lentiviral particle production, p24 concentration (in pg/mL) was quantified by ELISA according to kit instructions (Advanced Bioscience Laboratories Cat. #5421). All viral supernatants were measured in technical duplicate at a 1:100,000 dilution.

## Data availability

All code used for the above analyses is available at github.com/emmahodcroft/cluster_scripts (the commit tagged `journal_submission` was used to generate the figures in this manuscript). The code used to run the cluster builds is available at github.com/emmahodcroft/ncov cluster. Sequence data were obtained from GISAID and tables listing all accession numbers of sequences are available as supplementary information.

## SUPPLEMENTARY MATERIAL

### Observations for individual countries

There are multiple non-European samples in the cluster, from Hong Kong, Australia, New Zealand, and Singapore. All are likely exports from Europe, and indicate multiple introductions that were likely detected in quarantine on arrival. Interestingly, seven of the sequences from New Zealand appear to be linked to in-flight transmission en-route to New Zealand, likely originating from two passengers from Switzerland (Swadi et al., 2020). The observations below refer to sequences sampled before the end of September 2020 and are meant to highlight specific notable patterns.

### Norway

The first Norwegian samples, in July and August, seem likely to be a direct introduction from Spain, as they cluster tightly with Spanish sequences and the first sample (29th July) was just after quarantine-free travel to Spain was stopped. This introduction appears to have been contained, as no later-dated sequences descend from this group, and the number of 20E (EU1) sequences in Norway drops to zero by the end of August. However, 20E (EU1) sequences appear in Norway once again from mid-September and grew to around 20%, where it has remained steady (Fig 2). Unlike the initial sequences, these later sequences fall across the tree, and often cluster more closely with diversity in European countries other than Spain, which may suggest further introductions came from third countries (see 20E (EU1) Nextstrain build online).

### Switzerland

Quarantine-free travel to Spain was possible from 15th June to 10th August. The majority of holiday return travel is expected from mid-July to mid-August towards the end of school holidays. When all lineages circulating in Switzerland since 1 May are considered, the notable rise and expansion of 20E (EU1) is clear (see Fig S7).

To observe 31 genotypes observed in Switzerland that are also observed in another country prior to 2020-09-30, suggesting an introduction into Switzerland, directly from Spain or indirectly, through a third country. Three of the 31 nodes involve more than twenty Swiss sequences, and seem to have grown rapidly, consistent with the growth of the overall cluster.

For those nodes that don’t directly or through their parents share diversity with Spanish sequences, the Swiss sequences are most closely related to diversity found in the UK, France, and Denmark, suggesting possible transmission between other EU countries and Switzerland or diversity in Spain that was not sampled.

### Belgium

Along with many European countries, Belgium reopened to EU and Schengen Area countries on the 15th June. Belgium employed a regional approach to travel restrictions, meaning that while travellers returning from some regions of Spain were subject to quarantine from the 6th of August, it was not until the 4th September that most of Spain was subject to travel restrictions. Belgian sequences share diversity with sequences from Spain, the UK, Denmark, the Netherlands, and France, among others, and are spread across 22 nodes in the phylogeny.

### Netherlands

The Netherlands began imposing a quarantine on travellers returning from some regions of Spain on the 28th July, increasing the areas from which travellers must quarantine until the whole of Spain was included on the 25th August. 25 nodes across the phylogeny contain sequences from the Netherlands. On twelve nodes sequences from the Netherlands share diversity with Spanish sequences, suggesting possible direct importations from Spain. The earliest sample from the Netherlands was identified on the 20th June, the same date as the first sequences from Spain. However, travel began increasing from Spain to the Netherlands markedly earlier than to most European countries (Fig. 4 A), and this Dutch sequence nests within the diversity of early sequences from Spain, suggesting this sequence is the result of the earliest export of the variant outside of Spain.

### Denmark

Denmark re-opened their borders to the majority of European countries on the 27th of June. By the end of July, however, the government was advising travellers to Spain’s Aragon, Catalonia, and Navarra regions to be tested for SARS-CoV-2 on their return. On the 6th of August, Denmark advised against all non-essential travel to Spain, and strongly suggested quarantine on return, though notably quarantine has not been a legal requirement, as it has been in many other countries in Europe. The 977 sequences prior to Oct 2020 included in our analysis from Denmark are found on 44 nodes across the phylogeny.

### The UK and Ireland

The first sequences in the UK (Wales) which associate with the cluster are from the 7th July, just before the period from the 10th to 26th July when quarantine-free travel to Spain was allowed for England, Wales, and Northern Ireland. The first Irish sequences to associate with the cluster were taken a short time later, on the 23rd of July.

The large number of sequences from the United Kingdom make introductions harder to quantify. A total of 125 nodes in the phylogeny contain sequences from the United Kingdom. The remaining nodes most often share diversity with Denmark, Switzerland, and Ireland.

The 108 sequences of the 20E (EU1) variant from Ireland cluster in 17 nodes on the phylogeny. Notably, 15 of these 17 nodes containing Irish sequences also shares diversity with sequences from the United Kingdom. However, as mentioned before, the diversity in Spain is likely not fully represented in the tree, so direct transmission cannot be ruled out.

### Differing Travel Restrictions in the UK and Ireland

While quarantine-free travel was allowed in England, Wales, and Northern Ireland from the 10th–26th July, Scotland refrained from adding Spain to the list of ‘exception’ countries until the 23th July (meaning there were only 4 days during which returnees did not have to quarantine). On the other hand, Ireland never allowed quarantine-free travel to Spain, but did allow quarantine-free travel from Northern Ireland. Similarly, Scotland allowed quarantine-free travel to and from England, Wales, and Northern Ireland. Despite having only a very short or no period where quarantine-free travel was possible from Spain, both Scotland and Ireland have cases linked to the cluster consistent with significant travel volume between Spain and these countries over the summer. Additionally, close connections to the UK countries with similarly high travel volumes may have allowed further introductions.

**Figure S 1.**
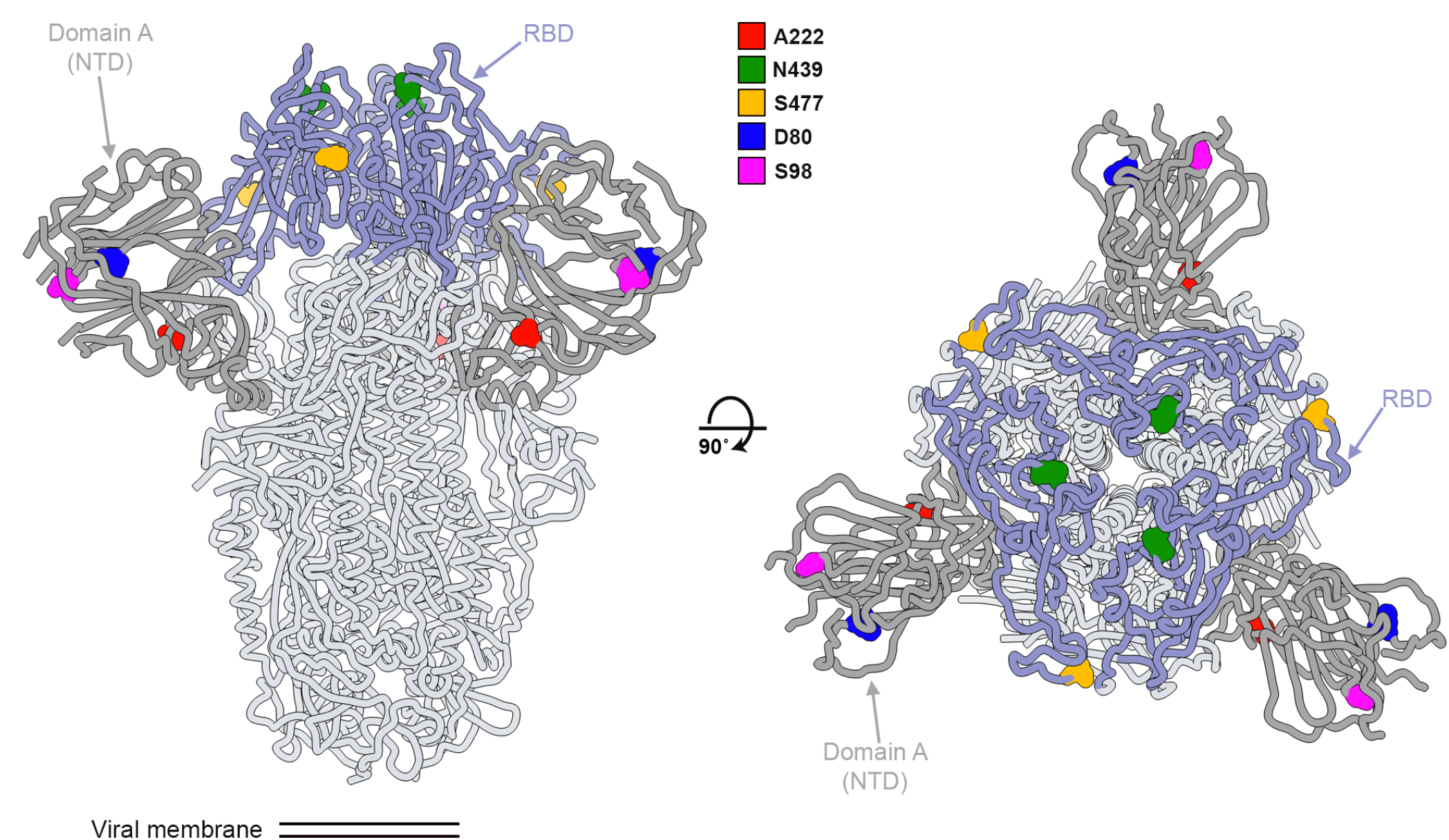
Two orthogonal orientations of the SARS-CoV-2 spike glycoprotein trimer highlighting the position of the variants described in the manuscript and the receptor binding domain (RBD) and the NTD (domain A). 222: red; 439: green; 477: orange; 80: blue; 98: magenta

**Figure S 2.**
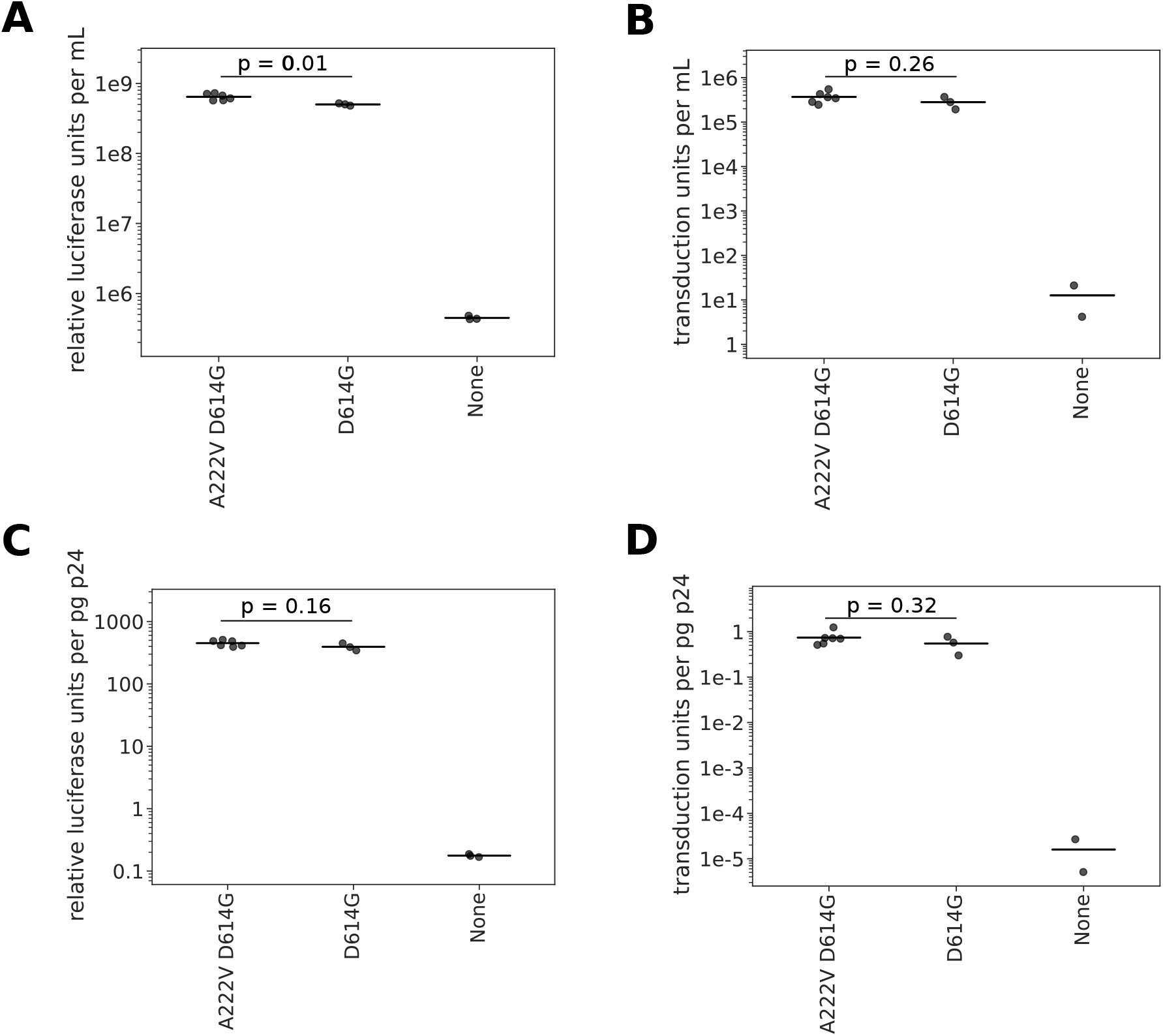
Titers of lentiviral particles pseudotyped with spike with or without the A222V mutation. A) Titers of lentiviral particles carrying luciferase in the viral genome. The horizontal line indicates the mean titer. B) Titers of lentiviral particles carrying the fluorescent protein ZsGreen in the viral genome. The horizontal line indicates the mean titer. In both cases, titers with the A222V mutation are on average higher by a factor 1.3. C) Titers of lentiviral particles carrying luciferase in the viral genome normalized by the p24 concentration (pg/mL) of each viral supernatant. After p24 normalization, the titer difference shrinks fom 1.28 to 1.14 fold, increasing the p-value to 0.16. D) Titers of lentiviral particles carrying ZsGreen in the viral genome normalized by the p24 concentration (pg/mL) of each viral supernatant. All *p*-values calculated using a two-sided *t*-test.

**Figure S 3.**
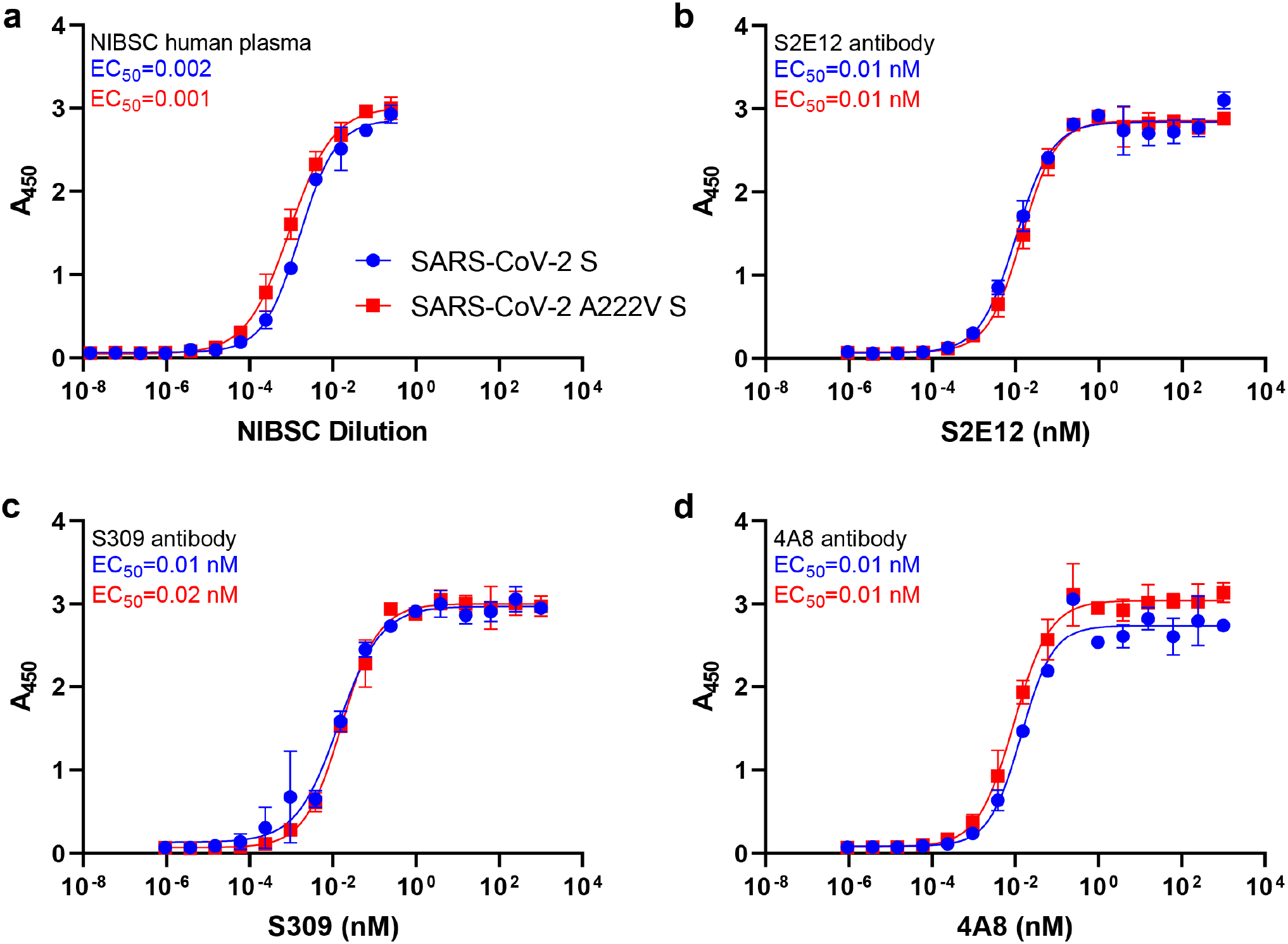
The substitution A222V in spike has no substantial effect on antigenic properties. (**a**) Binding of a serial dilution of NIBSC convalescent plasma to immobilized SARS-CoV-2 2P S (blue) or SARS-CoV-2 2P A222V D614G S (red). (**b-c**), Binding of serially diluted concentrations of the human neutralizing antibodies S309 (**b**) and S2E12 (**c**) to immobilized SARS-CoV-2 2P S (blue) or SARS-CoV-2 2P A222V D614G S (red). (**d**) Binding of serially diluted concentrations of the human neutralizing antibody 4A8 to immobilized SARS-CoV-2 2P S (blue) or SARS-CoV-2 2P A222V D614G S (red). *n* = 2 experiments performed with independent protein preparations (each in duplicate).

**Figure S 4.**
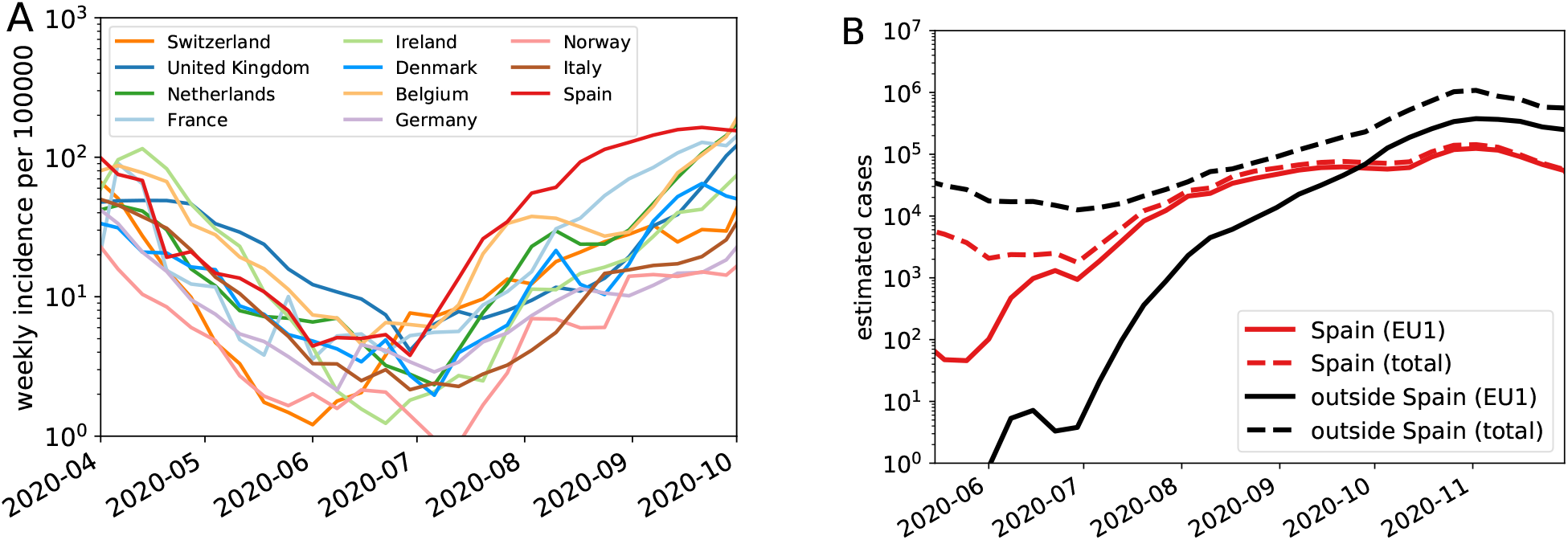
Incidence in various countries over the summer. **A**: Spain and Belgium had relatively higher incidence from the start of July compared with other countries in Europe. **B**: The estimated total number of EU1 cases (red) outside of Spain (countries as in A) surpasses the cases in Spain in September.

**Figure S 5.**
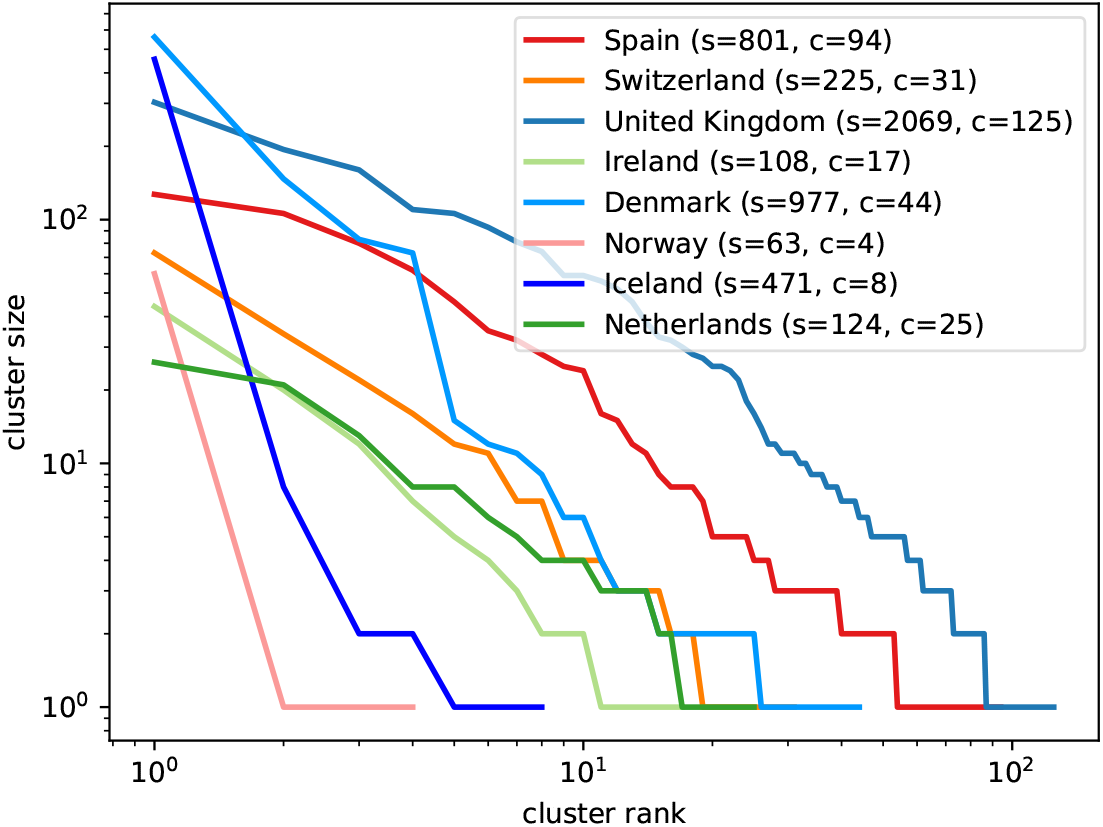
Rank-order plots of sizes of clusters of sequences in different countries compatible with a single introduction. Countries like Norway and Iceland have relatively few clusters, with one or two large clusters dominating, suggesting a small number of introductions dominated 20E (EU1) circulation. Countries like the UK and Denmark, on the other hand, show many clusters of varying size, indicating multiple introductions that led to onward spread. The legend indicates total number of sequences *s* and number of clusters *c*.

**Figure S 6.**
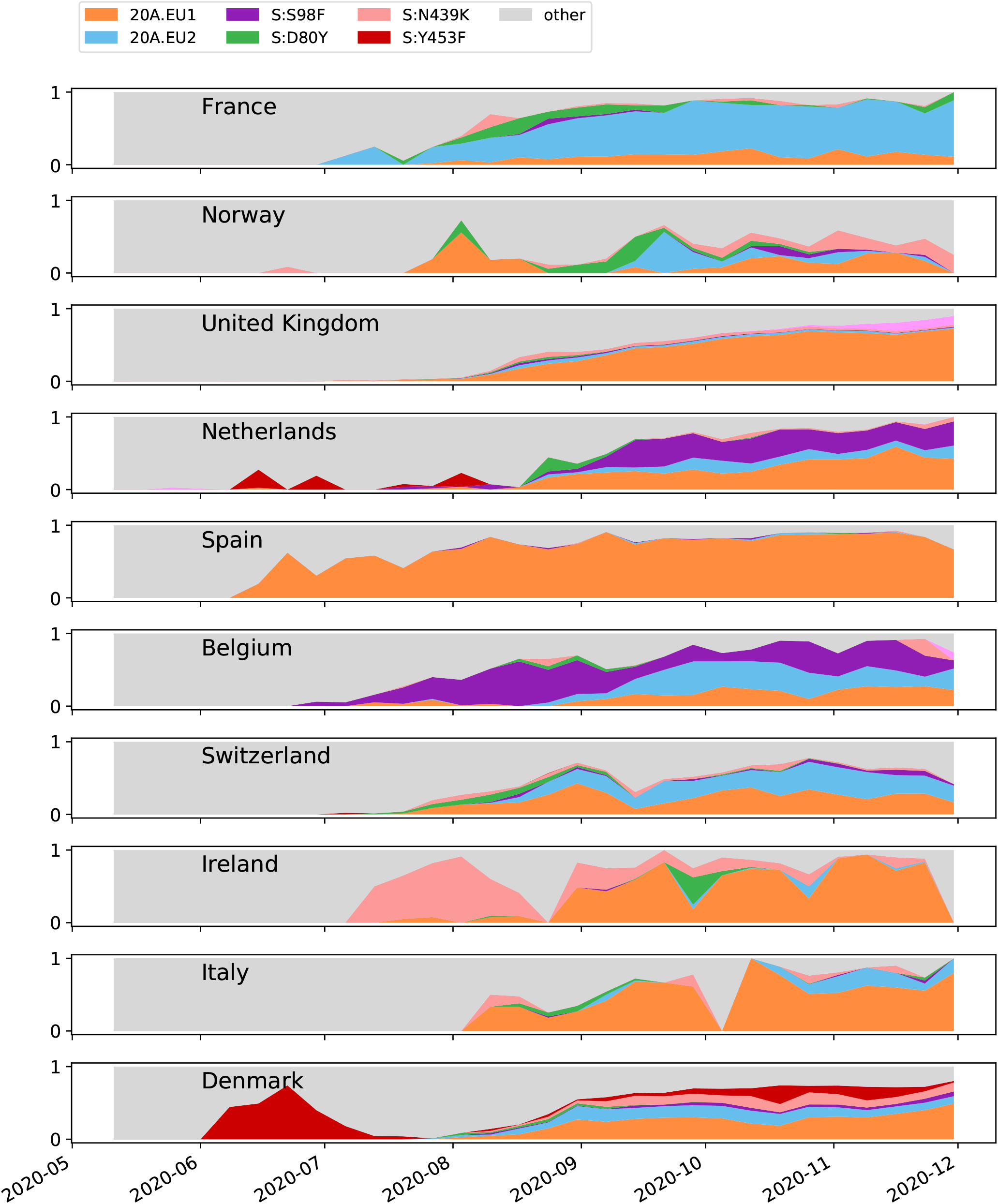
In countries with at least ten sequences that fall into any of the defined clusters, the proportion of sequences per ISO week that fall into each cluster is shown.

**Figure S 7.**
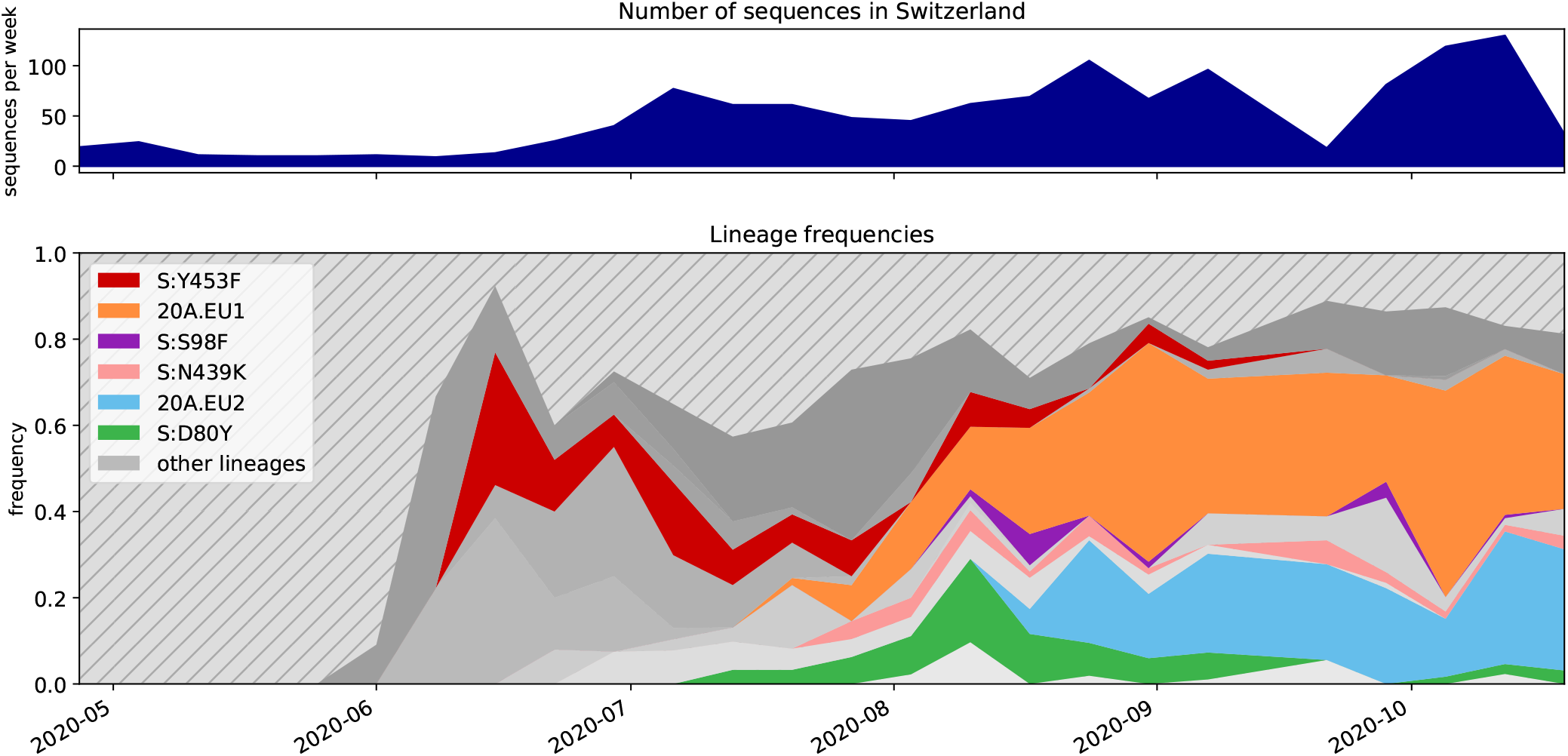
Lineages found in a Swiss-focused Nextstrain build. A lineage is defined as a node present in the tree after the cutoff date of 1 May 2020 with at least 10 Swiss sequences as children. Clusters discussed in this manuscript are labelled. Lineages are shown as the proportion of the total number of sequences per week in Switzerland. Striped space in the bottom graph represents lineages with most recent common ancestors dating back prior to 1 May 2020 and lineages that do not contain at least 10 Swiss sequences.

**Figure S 8.**
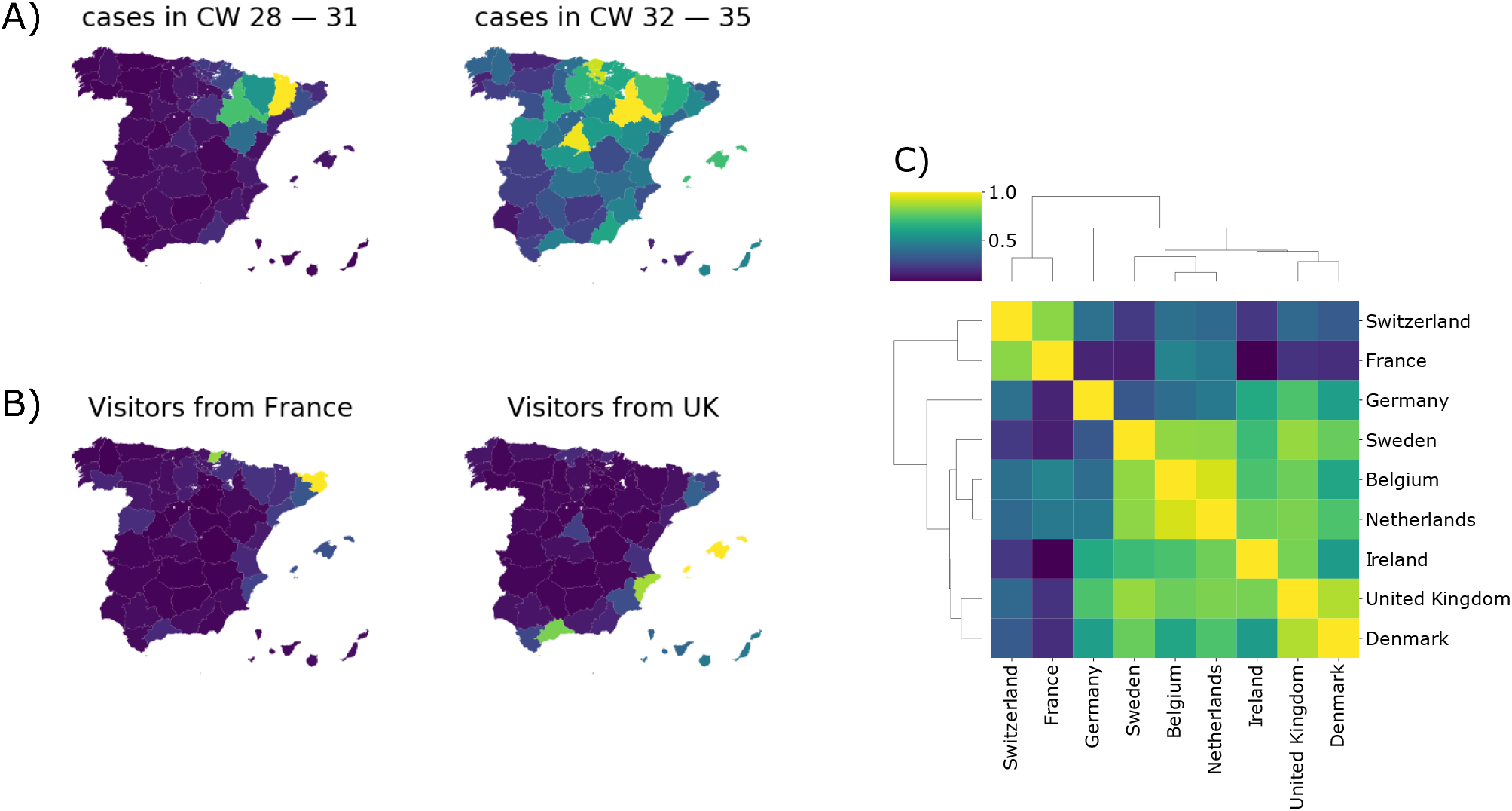
**A**: Incidence in Spain in early and mid-summer. **B**: Distributions of visitors from Spain from different countries. **C**: Similarities of destinations in Spain among visitors from different countries in calendar weeks 28-35.

**Figure S 9.**
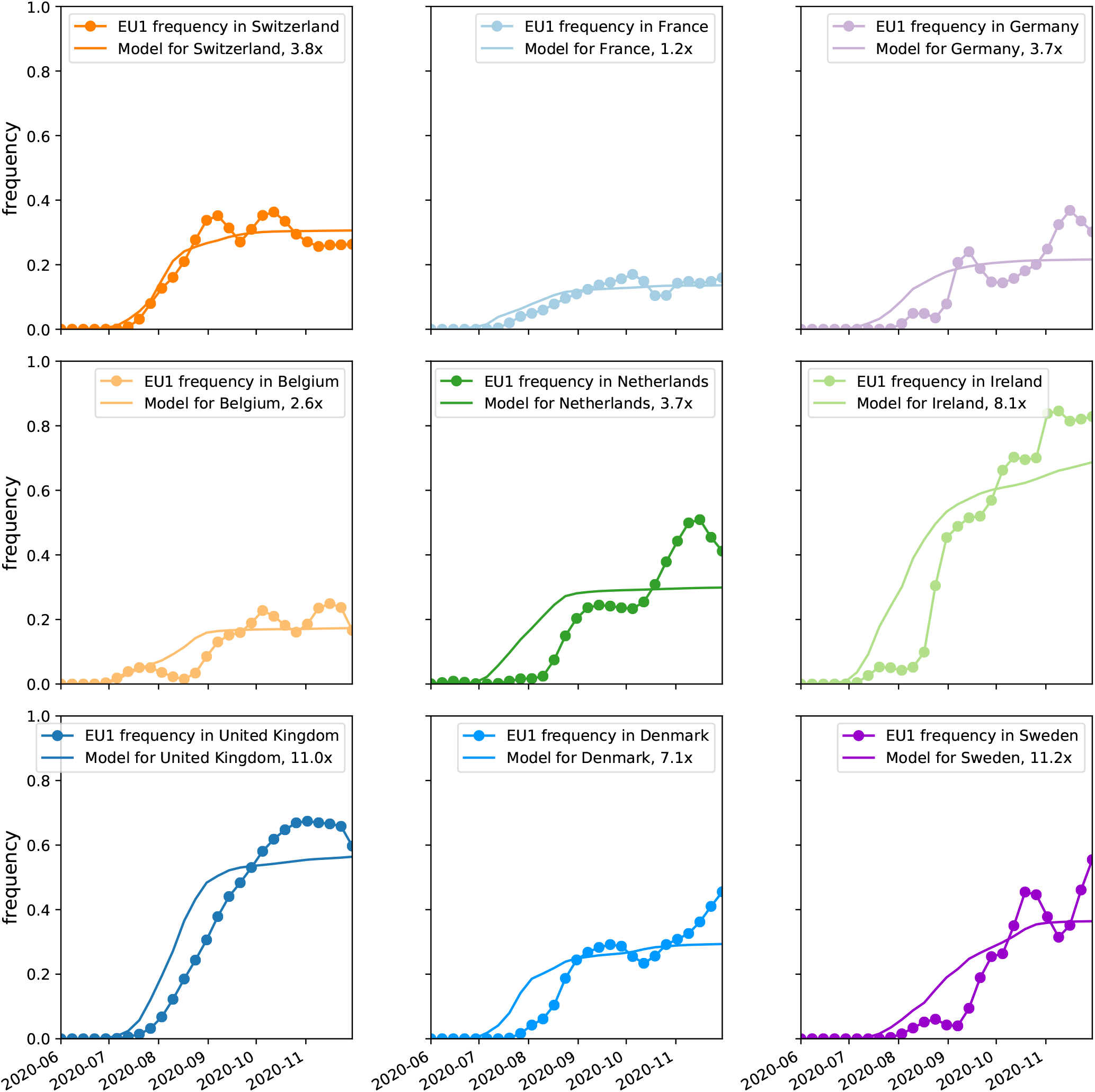
Rescaled predictions by the import model match observed frequency trajectories. In most countries, observations of 20E (EU1) increased in July 2020 and reached a plateau or a slower increase by Oct 2020. Predictions by the import model need to be scaled (see legend) to match the observed frequencies by a factor between 1.2 and 11 (see main text for discussion). Fluctuations on short time scales in the observed frequency of 20E (EU1) are likely due to sampling and dynamics of local outbreaks. Observed frequencies are subject to variable reporting delays.

**Figure S 10.**
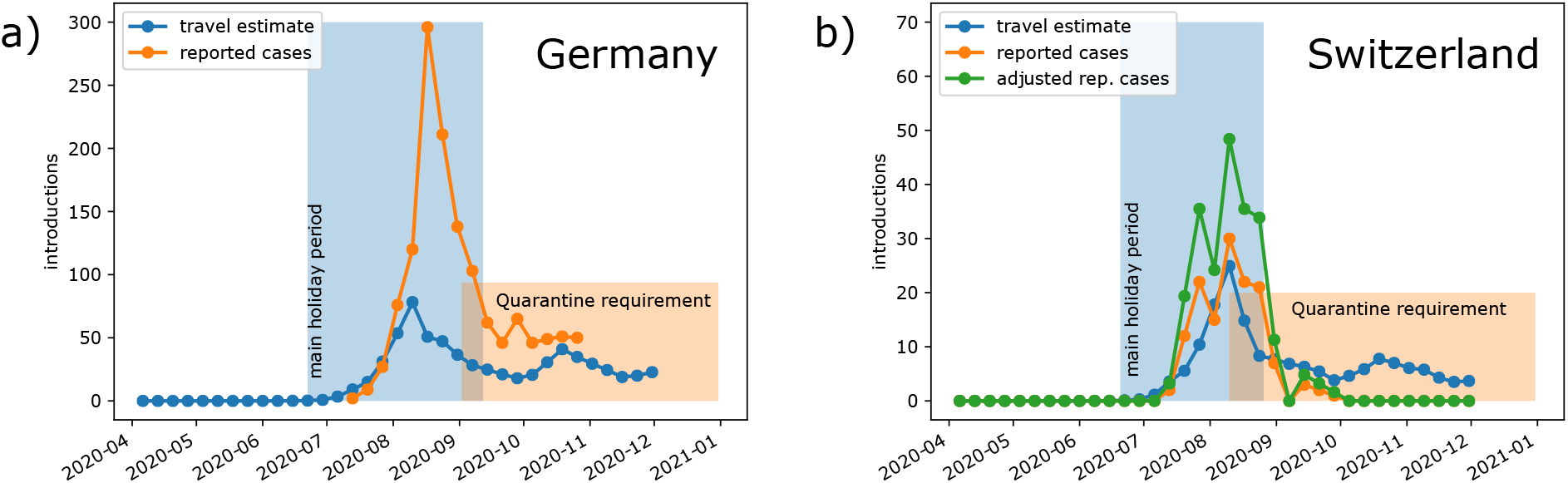
Reported and estimated introductions of 20E (EU1) to Germany (a) and Switzerland (b). **Travel estimate** is estimated introductions from Spain based on incidence and roaming data. **Reported cases** are cases with a suspected origin in Spain as reported by the RKI (Robert-Koch-Institute, 2020) and the Federal Office of Public Health (FOPH), for Germany and Switzerland, respectively. In Switzerland the **adjusted rep. cases** accounts for the fact that 37% of case reports lack exposure information.

**Figure S 11.**
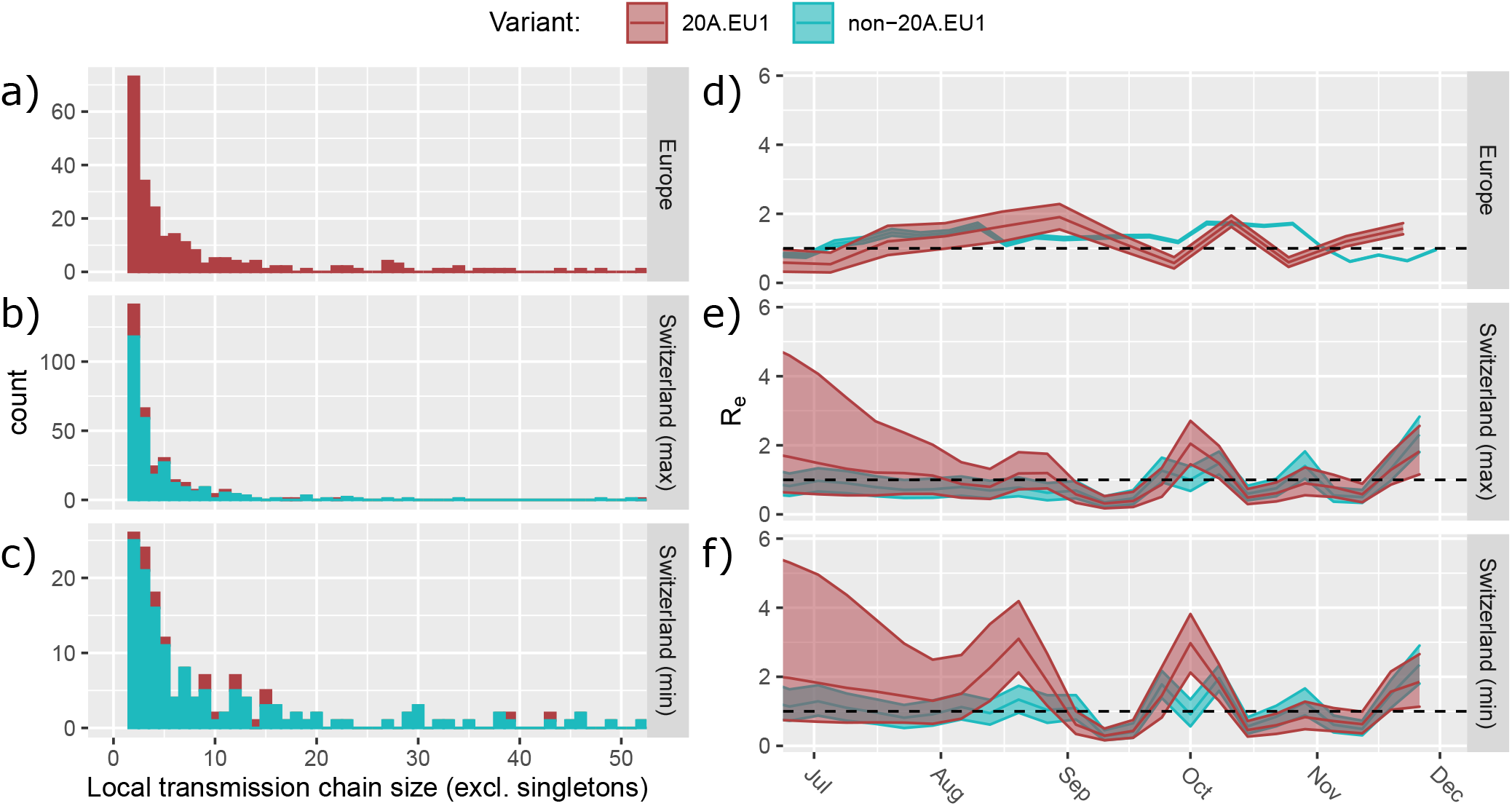
Introduction and spread of the 20E (EU1)-variant across Europe (top panels: a & d) and in Switzerland (bottom panels: b, c, e, f). **(a-c)** The size of putative transmission chains caused by introductions into Europe and Switzerland. Not shown are the number of singletons, which are introductions with no evidence of onward transmission. In Switzerland, these are shown under two extreme definitions of an introduction (min/max; see Methods). Depending on the min/max definition of introductions, there were between 14 or 236 singletons of 20E (EU1) (41 or 81% of all 20E (EU1) introductions) and 62 or 1089 non 20E (EU1) (30 or 79% of all non-20E (EU1) introductions). In Europe, we see 206 20E (EU1) singletons (46% of all 20E (EU1) introductions). There were also a small number of larger transmission chains including more than 53 transmissions (20 across all data sets) which are not shown in the histograms. **(d-e)** The effective reproductive number estimated for 20E (EU1) (red) and the non-20E (EU1) variants (blue). In Switzerland, this is done for the two extreme definitions of an introduction. For Europe, non-20E (EU1) *R*_*e*_ estimates were generated from case numbers. While there is little data to inform estimates of *R*_*e*_ for 20E (EU1) in July and it differs little from the prior, there is some evidence that 20E (EU1) was growing faster than other variants in August. However, systematic differences in ascertainment in travel associated cases might confound this inference. From mid-September, *R*_*e*_ of 20E (EU1) is largely statistically indistinguishable from that of other variants. Shaded areas indicate 95% HPD regions. Notably, the peak in August in the Swiss analysis is larger under the ‘min’ definition (f) than under the ‘max’ definition (e), consistent with a more conservative definition of a cluster which would then require more onward transmission.

**Figure S 12.**
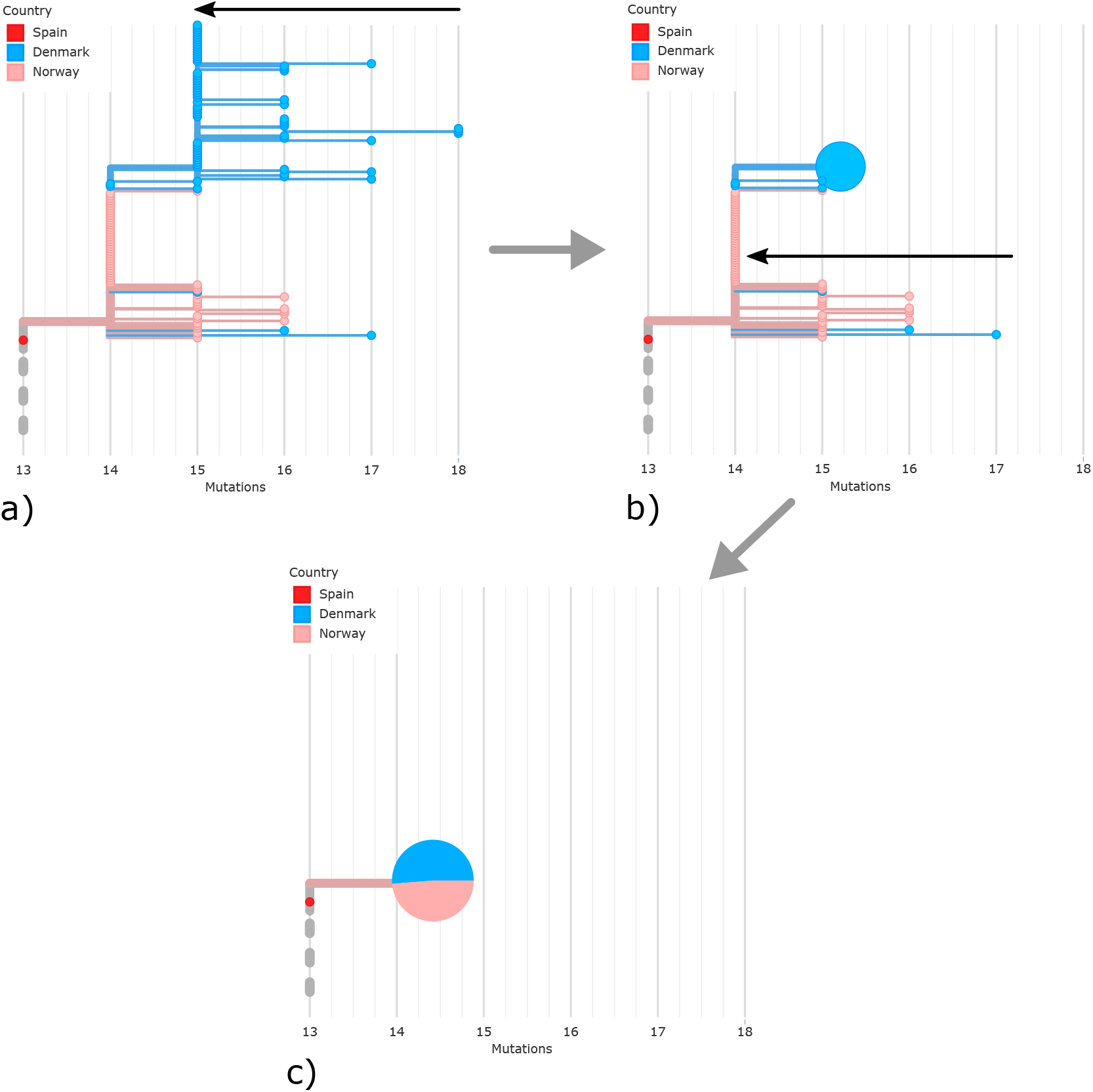
An example illustration of how the pie-chart phylogeny in Fig 3. The tree is shown in ‘divergence view’ with the branch lengths in mutations. Internal nodes are shown as horizontal lines with other nodes (internal and external) branching from them. If sequences are identical, they align on the horizontal line. In this example zooming in to the Norwegian cluster, the outermost tips are first collapsed down to their parental node (a), forming a pie chart that consists only of sequences from Denmark (b). This single-country pie chart is collapsed with the next level of nodes (c), including more sequences from Denmark and sequences from Norway, to form a multi-country pie chart.

**Table S I.** Summary of sequences observed in the 20E (EU1) cluster, sorted by the date of the first observed sequence. Data for analysis was cut off at the 30 November 2020 (see Methods).

| Country | First Observation | # Sequences | Last Observation |
| --- | --- | --- | --- |
| Spain | 2020-06-20 | 1442 | 2020-11-30 |
| Netherlands | 2020-06-20 | 577 | 2020-11-30 |
| United Kingdom | 2020-07-07 | 48251 | 2020-11-30 |
| Wales | 2020-07-07 | 7500 | 2020-11-30 |
| England | 2020-07-18 | 35627 | 2020-11-30 |
| Northern Ireland | 2020-07-21 | 440 | 2020-11-20 |
| Scotland | 2020-08-01 | 4538 | 2020-11-30 |
| Belgium | 2020-07-17 | 220 | 2020-11-30 |
| Switzerland | 2020-07-22 | 713 | 2020-11-30 |
| Ireland | 2020-07-23 | 402 | 2020-11-29 |
| Norway | 2020-07-29 | 149 | 2020-11-29 |
| France | 2020-08-01 | 141 | 2020-11-30 |
| Denmark | 2020-08-03 | 4654 | 2020-11-30 |
| Germany | 2020-08-07 | 179 | 2020-11-30 |
| Italy | 2020-08-13 | 333 | 2020-11-30 |
| Iceland | 2020-08-13 | 2430 | 2020-11-30 |
| Sweden | 2020-08-14 | 67 | 2020-11-25 |
| Hong Kong | 2020-08-15 | 4 | 2020-11-30 |
| USA | 2020-08-18 | 15 | 2020-11-30 |
| Australia | 2020-08-20 | 17 | 2020-11-29 |
| Lithuania | 2020-09-07 | 16 | 2020-11-19 |
| Luxembourg | 2020-09-21 | 324 | 2020-11-30 |
| New Zealand | 2020-09-22 | 27 | 2020-11-20 |
| Czech Republic | 2020-09-24 | 2 | 2020-11-25 |
| Singapore | 2020-10-14 | 8 | 2020-11-15 |
| Portugal | 2020-11-02 | 290 | 2020-11-24 |

**Table S II.**
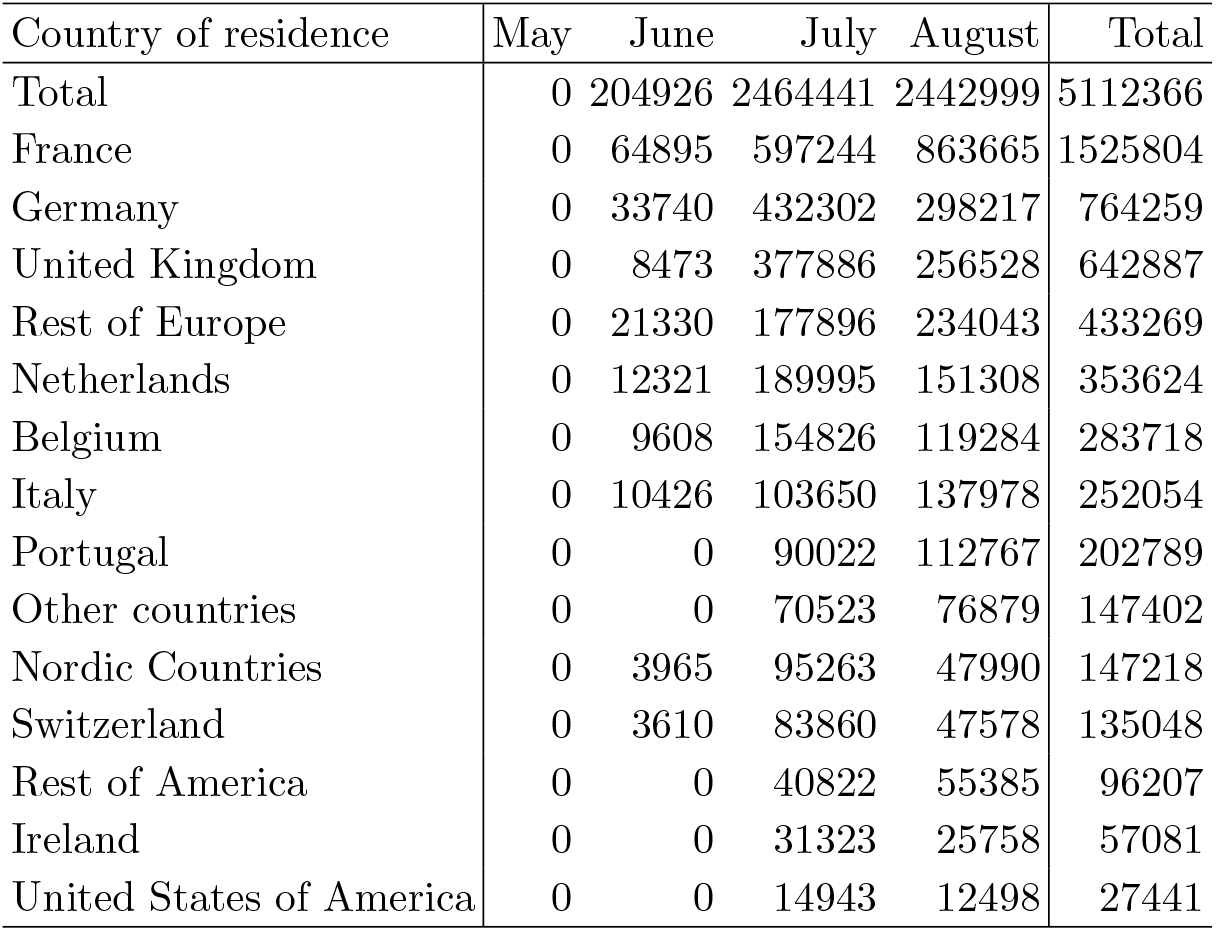
Arrival statistics of tourists in Spain over the Summer 2020 (Instituto Nacional de Estadistica, 2020)

